# *serojump*: A Bayesian tool for inferring infection timing and antibody kinetics from longitudinal serological data

**DOI:** 10.1101/2025.03.04.25323335

**Authors:** David Hodgson, James Hay, Sheikh Jarju, Dawda Jobe, Rhys Wenlock, Thushan I. de Silva, Adam J. Kucharski

## Abstract

Understanding acute infectious disease dynamics at individual and population levels is critical for informing public health preparedness and response. Serological assays, which measure a range of biomarkers relating to humoral immunity, can provide a valuable window into immune responses generated by past infections and vaccinations. However, traditional methods for interpreting serological data, such as binary seropositivity and seroconversion thresholds, often rely on heuristics that fail to account for individual variability in antibody kinetics and timing of infection, potentially leading to biased estimates of infection rates and post-exposure immune responses. To address these limitations, we developed ***serojump***, a novel probabilistic framework and software package that uses individual-level serological data to infer infection status, timing, and subsequent antibody kinetics. We validated ***serojump*** using simulated serological data and real-world SARS-CoV-2 datasets from The Gambia. In simulation studies, the model accurately recovered individual infection status, population-level antibody kinetics, and the relationship between biomarkers and immunity against infection, demonstrating robustness under observational noise. Benchmarking against standard serological heuristics in real-world data revealed that ***serojump*** achieves higher sensitivity in identifying infections, outperforming static threshold-based methods and precision in inferred infection timing. Application of ***serojump*** to longitudinal SARS-CoV-2 serological data taken during the Delta wave provided additional insights into i) missed infections based on sub-threshold rises in antibody level and ii) antibody responses to multiple biomarkers post-vaccination and infection. Our findings highlight the utility of ***serojump*** as a pathogen-agnostic, flexible tool for serological inference, enabling deeper insights into infection dynamics, immune responses, and correlates of protection. The open-source framework offers researchers a platform for extracting information from serological datasets, with potential applications across various infectious diseases and study designs.

**AUTHOR SUMMARY:** Tracking how infections spread and how our immune systems respond to them is essential for improving public health. One way to study this is by analysing blood samples to measure antibody levels, which can help estimate who has been infected, when they were infected, and how their immune response has developed over time. However, traditional methods for interpreting these antibody levels often use simple cutoffs that do not account for how much people’s immune responses can vary, which can lead to inaccurate results. To solve this, we created a new tool in R called ***serojump***. This tool uses advanced statistical methods to analyse changes in antibody levels over time, helping to more accurately identify who has been infected, when they were infected, and what happens to their antibodies afterwards. In our tests, ***serojump*** was better at detecting infections than traditional methods and provided more detailed insights about how people’s immune systems responded to the virus and vaccines. It also revealed infections that were missed by standard testing methods. Our tool is flexible and can be used for many different diseases. By helping researchers and public health workers better understand infection patterns and immune responses, ***serojump*** can support efforts to control the spread of diseases and develop more effective treatments and vaccines.

## 1. INTRODUCTION

Serological samples can be analysed to detect the presence of biomarkers, such as antibodies, made in response to an infection long after the infection has cleared.[1] By combining validated biomarker assays with appropriate statistical inference techniques, it is increasingly possible to estimate incidence rates, antibody kinetics, and population-level susceptibility without ongoing syndromic surveillance.[2] Different types of serological assays contribute to such analyses, including morphological assays such as ELISA, which measure antibody concentrations, and functional assays such as neutralisation tests, which measure the ability of antibodies to inhibit infection in vivo. Analysing serological samples is important because syndromic surveillance systems typically only capture cases of disease, representing the upper parts of the reporting pyramid.[3] However, effective control and prediction of pathogen spread require understanding the full spectrum of the underlying epidemiology, including both symptomatic and asymptomatic infections, as asymptomatic or pauci-symptomatic individuals may also contribute to transmission dynamics and population immunity. Therefore, analysing serological samples can enable researchers and healthcare professionals to infer crucial information about the epidemiology of a pathogen at the individual and population level, which case-based surveillance systems may otherwise miss.[4] Such analysis can in turn help the understanding of the immune system’s ability to combat various pathogens, aid in developing new targeted intervention programmes and provide insights into patterns and drivers of the transmission dynamics of infectious diseases.

On the individual level, past infection with a specific pathogen has traditionally been inferred from measured antibodies using either i) an antibody threshold level (i.e. seropositivity) or ii) a threshold fold-rise between a pair of samples (i.e. seroconversion).[3] Considerable research has focused on understanding how seropositivity and seroconversion rates change according to controlled host factors, such as age, geography, living conditions, sexual behaviour, etc.[5–7] On the population level, serological samples which are representative of a population (e.g. cross-sectional samples) can be used to estimate the prevalence of infectious diseases (seroprevalence) and determine how seroprevalence changes over time according to host factors.[8–10]

Estimation of infection through analysis of seropositivity or seroconversion requires deriving an accurate absolute or relative threshold value, and these are often determined by rule-of-thumb heuristics (e.g. for influenza: 4-fold-rise for conversion, titre of 1:40 HAI for seropositivity).[11] However, antibody responses vary greatly between individuals for many pathogens. Therefore, relying on these pre-determined heuristics to determine infections in serology studies can lead to both false positives and false negatives in inferred infection status, leading to biased estimates of prevalence.[8,12,13]) Consequently, there has been a growth in new analytical methods to make better use of quantitative measurements derived from serological samples to inform infectious disease epidemiology and public health policy, a field of research termed ’serodynamics’.[2]

In particular, efforts have been made to probabilistically infer individuals’ infection status and timing in a serological cohort by analysing longitudinal changes in serological titre.[14–16] By modelling the expected antibody level over time following infection or vaccination, single or multiple measurements of an individual’s antibody level can be used to back-calculate the presence of and likely timing of infection. These methods of inferring infection using an individual’s changes in antibody levels are called “time-since-infection” or “titre-based” methods. However, the complexity of modelling these uncertain parameters—such as antibody kinetics, individual infection status, and the timing of infection—requires sophisticated statistical methods. Standard Bayesian inference techniques, such as the Metropolis-Hasting algorithm, cannot sample over the high-dimensional, transdimensional space these models create, particularly when infection status and timing are being inferred.

One promising approach is reversible jump MCMC, a transdimensional sampler that can explore infection status and timing on a continuous scale.[17] This method has already provided valuable insights in specific studies, revealing patterns of infection and immunity for dengue and influenza that would otherwise remain hidden.[15,16] However, its application has been limited to narrowly defined problems due to the complexity of its underlying statistical methods and the need for problem-specific customisation of the sampler itself. This restriction prevents the broader adoption of reversible jump MCMC in epidemiological research despite its potential to infer accurate estimates of infection risk, correlates of protection, and antibody kinetics using only serological data.

Therefore, our study introduces *serojump*, a general-purpose statistical framework that applies reversible jump MCMC to infer infection status, timing, and antibody kinetics from serological data. By leveraging individual-level longitudinal changes in biomarker values, *serojump* provides a probabilistic approach to estimating epidemiological parameters, addressing the limitations of static serological heuristics. We validate this framework through simulation studies and real-world SARS-CoV-2 serological data from The Gambia, demonstrating its ability to recover infection histories, improve sensitivity in infection detection compared to traditional methods, and infer correlates of protection. By making these advanced inference techniques accessible via an R package, *serojump* enhances the utility of serological data for infectious disease surveillance and public health decision-making.

## 2. METHODS

### 2.1. Overview of inference framework

Using data on individual-level temporal changes in serological biomarker values, *serojump* can infer several key quantities within a single probabilistic framework: i) the mean population-level kinetics of the biomarker; ii) the individual-level probability of infection during a specific period; and iii) the distribution of timing of this infection. Defining as the measured value at time,, to biomarker, for individual ; as the latent binary vector representing the infection status of each individual (1: infected, 0: not infected), and = {τ_i_} is the infection time for those infected (where the length of the vector is equal to the number of infected individuals) we define the likelihood of this algorithm as in Menezes et al:[18]:

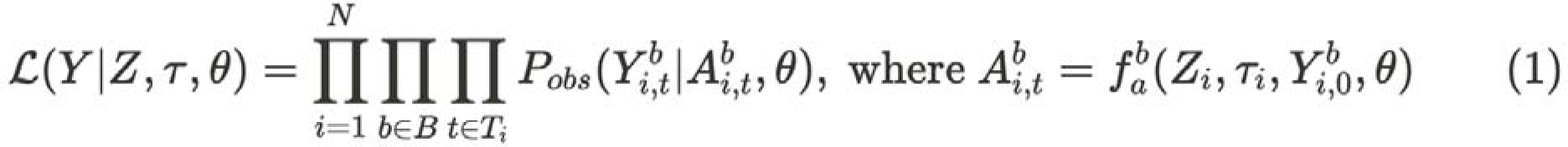

Where, is the observational model, describing the likelihood of the serological data,, given the model predicted latent titre values,, (defined by the function,) and represents the model parameters being estimated.

The posterior distribution is therefore defined by:

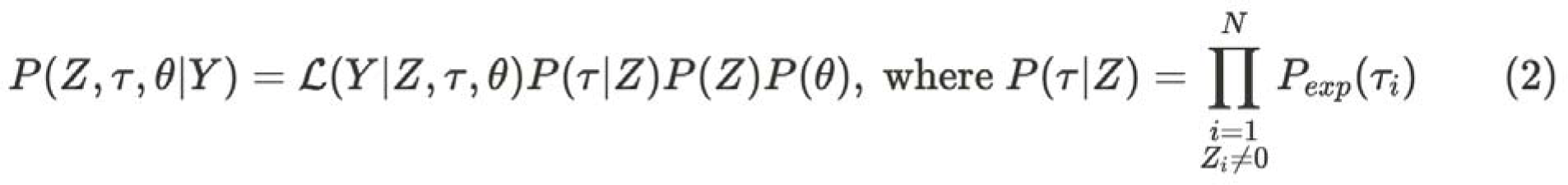

Where is the prior distribution for the timing of infection for individual *i*, is the prior distribution on the infection status binary vector, and is the prior distribution on the parameters describing the observational model and antibody kinetics model. To sample from EQUATION 2, we used a reversible-jump MCMC algorithm, which allows for efficient exploration over the fixed dimensions of and the dynamically changing dimensions of The full algorithm and its derivation are given in **Supplementary Methods section 1.1–1.3**.

#### 2.1.1 Antibody kinetics model

In the most general case, the antibody kinetics model, for an individual *i*, we define *k* immunological stimuli, {e_1_, e_2_, …, e_k-1_, e_k_} which occur at times {t_e1,_ t_e2_, … t_ek-1,_ t_ek_}, such that t_ei_ < t_ei+1_ for all i. These could represent stimuli such as exposure to an infection or vaccine. If the functions f^b^_e1_, f^b^_e2_, …, f^b^_ek-1_, and f^b^_ek_ represent the antibody kinetics following immunological stimulus e_i_ for biomarker *b*, then, given a serological sample at time t_ek_ < *t\** < t_ek+1,_ we estimate the biomarker value (e.g. log neutralising titre) at this time A_i, t_^b^ in the model as:

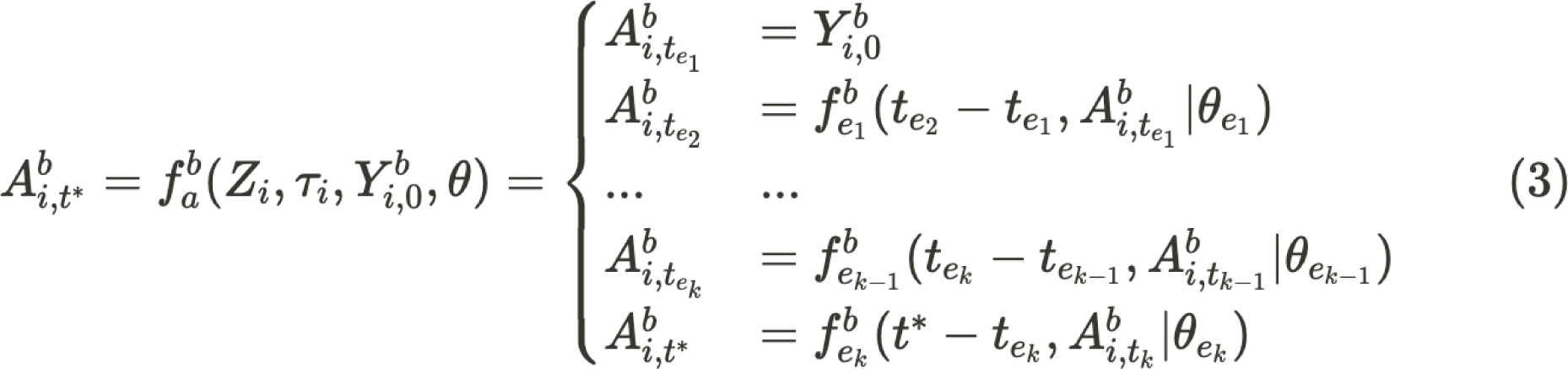

That is, the antibody titre is governed by an individual’s most recent exposure and the function for this response is conditional on the time since this exposure, the biomarker level at that time, and any exposure-specific parameters. Given that our analysis focuses on inferring infections, we assume that for a subset of individuals, the immunological stimuli, e_j_, results in an infection, and the timing of this infection r_i_ is t_ej_. We refer to this framework as an antibody kinetics model as, in this study, we are measuring titre values to antibody levels, but note the same method can be applied to any measured immunological biomarker (e.g. memory B cell concentrations).

#### 2.1.2 Observational model

The observation model defines the likelihood function that links the model-estimated biomarker value at the sampled time to the measured value at that time. Typically, serological measurements are assumed to follow a normal distribution: 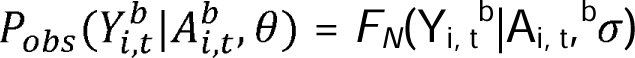. However, because all assays have upper and lower limits of detection, a censored normal distribution is often used to account for interval censoring. Additionally, many serological data points are discrete due to measurement constraints—such as dilution series in assays (e.g., 1:10, 1:20, 1:40, etc.)—where the true biomarker level is continuous, but observations are recorded in discrete steps. In such cases, a likelihood function that explicitly models interval-censored or discretized normal distributions is required to accurately capture the measurement process.[19]

#### 2.1.3 Infection model

The prior for the infection time,, for each individual can be estimated on the population-level from existing serological surveys or surveillance data to identify likely periods of infection. For example, surveillance data can still indicate the distribution of infection burden over time, if not the true cumulative number of infections. A key feature of our framework is that can be specified to reflect a shared force of infection (FOI) across individuals, ensuring that infection times are conditionally dependent given the population-level FOI. This avoids the naive assumption that values are fully independent for each individual. If this prior is not specified in the model, or there is no known data to infer the epidemic curve, the model will assume a uniform risk of infection over the study period (**Supplementary Methods section 1.4)**.

#### 2.1.4 Prior distributions

The prior distribution on the parameters describing the observational model and antibody kinetics model, can be defined by the user and are assumed to be independent. The prior distribution on the infection status binary vector, as with similar inference problems,[19] has an implicit prior resulting from the definition of the reversible jump algorithm. This is Bin(0.5, N), or 50% of the population infected on average. To remove this implicit prior and instead assign a uniform distribution to the total number of infected individuals during the epidemic, we choose the combinatoric prior:

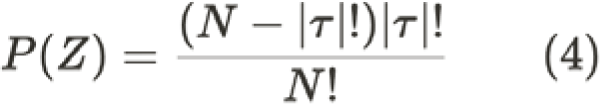

If prior knowledge on the number of missed infected individuals is known (e.g. from random community testing regardless of symptoms), these can be added to this prior (**Supplementary Methods section 1.4–1.5).**

#### 2.1.5. Post-processing of posterior distributions and correlates of protection

After fitting the *serojump* algorithm, we obtain posterior distributions for the set (τ̂, *Ẑ*, ^θ̂^). We calculate the posterior distribution of the post-exposure antibody trajectories for exposure *e* and biomarker *b, f^b^_e_*(*t*, ^θ̂^). To assess the posterior distribution of the number of infected individuals across the population, we calculate 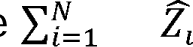 for each sample of the posterior distribution.A key estimate in serological studies is the relationship between a biomarker (e.g., antibody titre) and infection risk, where the biomarker is referred to as a *correlate of protection (COP)*. A COP represents either a direct mediator of immunity or a proxy for immune protection. To estimate this relationship from *serojump*, we fit a logistic curve with parameters *k* (gradient) and *x_0_*(mid-point) to describe how reference biomarker values relate to the probability of infection, given by:

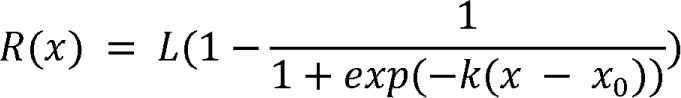

From this, we define the *absolute correlate of protection (aCOP)* as the fitted probability curve, representing the direct relationship between biomarker levels and protection from infection. It can be derived from the fitted R(x) as:

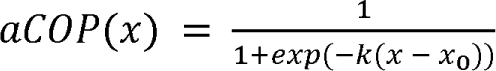

The *relative correlate of protection (rCOP)*, is a rescaled version of the aCOP that normalizes protection relative to a reference titre level (in this cas the lower limit of detection. It can also be derived from fitted R(x) as

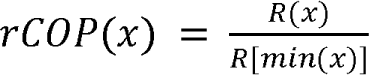

These estimates allow us to quantify the protective effect of immune markers and assess how biomarker levels modulate infection risk across a population (**Supplementary Methods section 1.6–7**).

### 2.2 Application to simulated data

#### 2.2.1 Description of the simulated data

To test whether it is possible to correctly recover epidemiological and immunological dynamics from common serological cohort structures, we first simulate a serological dataset using the *serosim* R package[18] to test the *serojump* framework. We simulate continuous epidemic serosurveillence (CES) cohort data, which represents a serostudy in which individuals are followed over an epidemic wave and sampled at multiple random time points throughout. The simulated data includes N = 200 individuals with serological samples taken within the first seven days of the study’s starting and a sample within the last seven days of the study’s ending. These individuals also had three samples taken randomly throughout the study during an 120-day epidemic wave.

We model the epidemic as a stochastic transmission process, where individuals can be either susceptible, exposed, or infected. The probability of exposure is set at 60% per individual, meaning that individuals coming into contact with an infectious person have this probability of becoming exposed, assuming no prior immunity. Among those exposed, the probability of progressing to infection is 30%, meaning that some individuals remain uninfected despite exposure. This corresponds to a scenario where a pathogen spreads within a population that starts with partial immunity. Over the study timeframe we assume that individuals can have a maximum of one exposure. To model a symmetric epidemic peak, we simulate the exposure time from a normal distribution, *N*(60, 20) days. To assess the impact of biomarker levels on infection risk, we simulate two different relationships between the measured biomarker and the probability of infection given exposure: (1) No COP model (Model A), where infection occurs with a fixed probability of 50% regardless of biomarker titre, representing a **non-titre-dependent correlate of protection**; and (2) With COP model (Model B), where infection probability follows a logistic function of biomarker titre, a commonly used correlate of protection model [20,21]. Explicitly, the simulated probability of infection, given a titre value, x, is given by f(x) = 1 / 1 +exp(−(−4 +2x)).

Following infection, the antibody kinetics are assumed to follow a linear rise to a peak at 14 days, followed by an exponential decay to a set-point value.[22] The formula for this biphasic trajectory is given by

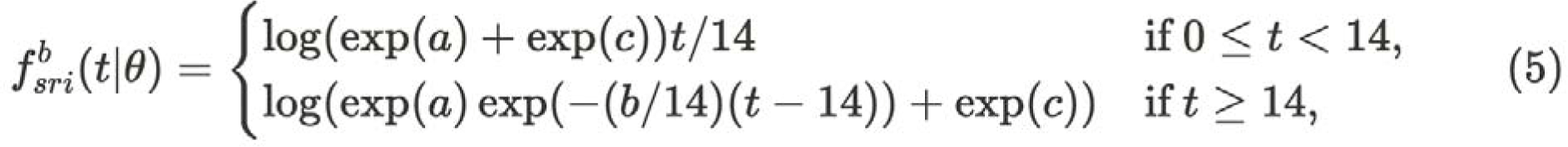

The simulated values we use are a = 1.5, b = 0.3, c = 1 and *t*, is the number of days post-infection. These values are calibrated to produce a biphasic response where antibodies **rise sharply** during the first 14 days post-infection (representing immune activation), followed by a gradual **ex**ponential decline after peak response, stabilising at a long-term set point. This pattern is commonly observed in infections such as influenza and SARS-CoV-2. If an individual is not infected over the period, their titre remains unchanged and, therefore, equal to their start titre throughout. We assume all individuals have the same antibody kinetic response following infection but we add observational noise into our model, assuming a normal distribution with standard deviation σ. In the base case, we assume σ = 0.1. We tested the robustness of the simulation recovery to noise by fitting the *serojump* algorithm to simulated data with increasing observational error. Specifically, we considered standard deviations in the normally distribution observation error for 11 values (0.01, 0.05, 0.1, 0.15, .., 0.45, 0.5), and evaluated the capacity for *serojump* to recover i) the infection status of individuals, ii) the timing of the epidemic and iii) the recovered population-level antibody kinetics.

#### 2.2.2 Model specification for *serojump*

We assume each individual’s serological profile is characterised by the **IgG biomarker** (b={IgG}). Two immunological events are considered: *e_pre_*, the pre-infection state and *e_inf_*, the infected state. The structural assumptions on the kinetics of the pre-infection and infection state are; for the pre-infection state, we assume the prior on the antibody levels Y_i_ are modeled as a linear wane

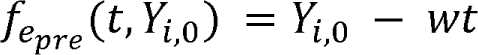

with a waning rate with the uniform distribution with prior *w* ∼ U(0,0.1). For the post-infection dynamics, we assume that antibody-kinetics boost according to the functional definition defined as 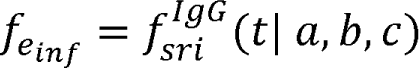 (Equation 5) with the priors, a ∼ *N*(2, 2), b ∼*N*(0.3, 0.05), c ∼ *U*(0, 4).

For the observational model, we assume that antibody measurements follow the normal distribution probability density function [*F_N_* (*X*|µ, σ)];

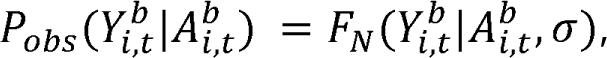

with the prior a ∼ U(0, 4). For the prior on the risk of infection over time, we assume this is uniform across the whole time period *P_exp_*(τ*_i_*) = *U*(1, 120) for all *i*. Finally, the distribution for the number of infection events, *P*(*Z*), is constrained by a combinatorial factor reflecting the arrangement of events across the population as described in Equation 4. A summary for the method specification for *serojump* can be found in the **Supplementary Methods Section 2**.

### 2.3 Application to empirical data

#### 2.3.1 Description of the real-world serological data on SARS-CoV-2 from The Gambia

To test *serojump* on real-world serological data, we used longitudinal serological data collected before and after the Delta wave [23] in a household cohort study of 349 individuals in The Gambia. Quantitative ancestral anti-spike and nucleocapsid (NCP) IgG data were generated using previously validated ELISA assays,[10] calibrated to the WHO International Standard for anti-SARS-CoV-2 immunoglobulin (cat no NIBSC 20/136). In addition to serological data, SARS-CoV-2 PCR testing was conducted weekly regardless of symptoms and additionally for those who presented with influenza-like illness symptoms during this period. In addition to PCR-positive diagnoses, dates of SARS-CoV-2 vaccination and PCR-positive pre-Delta infections were also known. At the start of the study, which was just prior to the Delta variant circulating, 56.7% of people were seropositive for SARS-CoV-2.[23] Over the study period, the Delta variant predominantly circulated with 99 PCR-confirmed cases, with an additional 21 PCR confirmed SARS-CoV-2 infections with pre-Delta variants, and 49 individuals who received a dose of vaccine. A description of the timing of the sample collection, and infections for each individual is given in **Supplementary Methods Section 3**.

#### 2.3.2 Model specification for serojump

Each individual’s serological profile is characterized by two biomarkers: IgG to SARS-CoV-2 ancestral spike and NCP protein (b={spike, NCP}). **We model** four immunological events: e_pre_, the pre-infection state, e_delta_, the Delta-infected state, e_pre-delta,_ the pre-Delta infection state, and e_vax_ the vaccination state. For the preinfection state, we assume the prior on the antibody levels Y_i_ for biomarker *b* are modeled as a linear wane:

with a waning rate, *w*, with a prior *w ∼ U(0, 0.1).* This parameter is estimated separately for each biomarker.. For post-infection (pre-Delta and Delta) and vaccination, we assume that antibody kinetics follow a **boosting function** as described in Teunis et al.[24] () and that the level of boosting is scaled based on the **l**og-transformed titre values at exposure (such that, where

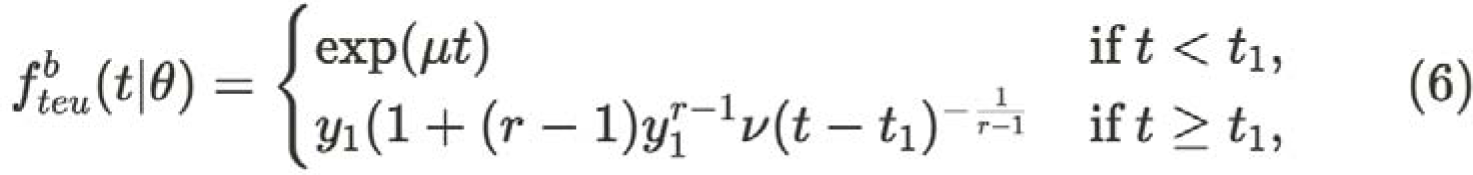

and

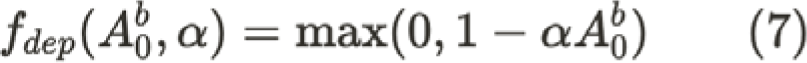

Where, v = 0.001, and the priors governing the boosting response are given by: r

∼ U(0, 1), y_1_ ∼ U(0, 6), t_1_ ∼ U(7, 21), and ∼U(0, 1). Thus, in the posterior distribution, each immunological event (pre-Delta infection, Delta infection, vaccination) and each biomarker has distinct parameter values describing its kinetics, but they share the same prior distributions.. For the observational model, we assume that antibody measurements follow the normal distribution probability density ;

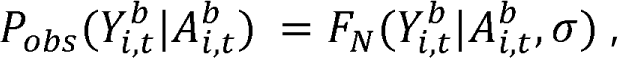

with prior a ∼ Exponential(1). We choose an empirical distribution based on the PCR positive data for the prior on the risk of infection over time. Finally, the distribution of a number of infection events, *P*(*Z*), is constrained by a combinatorial factor reflecting the arrangement of events across the population as described in Equation 4. A summary for the method specification for *serojump* can be found in the **Supplementary Methods Section 3**.

#### 2.3.3 Benchmarking against standard serological heuristics

We evaluate how well serojump and other serological approaches can infer true infection status (as defined by PCR-positivity) using serological data alone, without incorporating PCR results into the inference process. Specifically, we compare five different infection classification strategies based on serology: serojump, a four-fold rise in spike or NCP, and seropositivity thresholds for spike and NCP (IC50 values of 6.39 and 3.00) [23]. To assess the performance of these classification approaches, we compute and compare their sensitivity in detecting PCR-confirmed infections.

### 2.4. Implementation

The code is written in R (v.4.4.1)[25] and Rcpp (v1.0.14)[26], open source and packaged at https://github.com/seroanalytics/serojump. The package is flexible, allowing the user to input their functional forms and associated prior distributions for the antibody kinetics model (f ^b^) for each immunological stimulus and biomarker, or the user can select built-in functions from previous papers such as those mentioned in this study. Further, the user can specify their functional form for the observational model likelihood [P_obs_ (^b^ |A^b^)] and the prior distributions on the arguments. Finally, the user can customise the prior on the risk of infection over the time period, P_exp_(r_i_), and the prior on the distribution of infection events P(Z). The inputs for *serojump* to run the model on the simulated data and empirical data are summarised in T**ables S1–4**. The package *serojump* has vignettes to reproduce this study, which can be found at https://seroanalytics.org/serojump/articles/. The assessment of convergence statistics is summarised in **Supplementary Methods section 1.8**.

## 3. RESULTS

### 3.1 CASE 1: SIMULATED DATA

#### 3.1.1. Simulation recovery for the base case simulated dataset

For the default simulated dataset with low observational noise (Cf = 0.1), the individual-level antibody kinetics show good agreement between simulated and recovered kinetics under the With COP and No COP simulated data (Figure 1A–B) with the observed data points (grey dots) close to the median posterior distributions of the model-fitted individual-level antibody trajectories (red lines). We recover the infection status of each individual with 100% sensitivity and specificity, and the differences between simulated infection days and model-predicted infection days (Figure 1C–D) are small (within 14 days) for almost all individuals. This results in a close alignment between the simulated and the recovered epidemic curves (Figures 1E–F). These observations suggest that for the default simulated dataset, both the With COP and No COP data are well recovered, as evidenced by closer alignment of posterior medians to observed data, with minor errors in infection time predictions and an accurate reconstruction of the infection time distribution.

**Figure 1:**
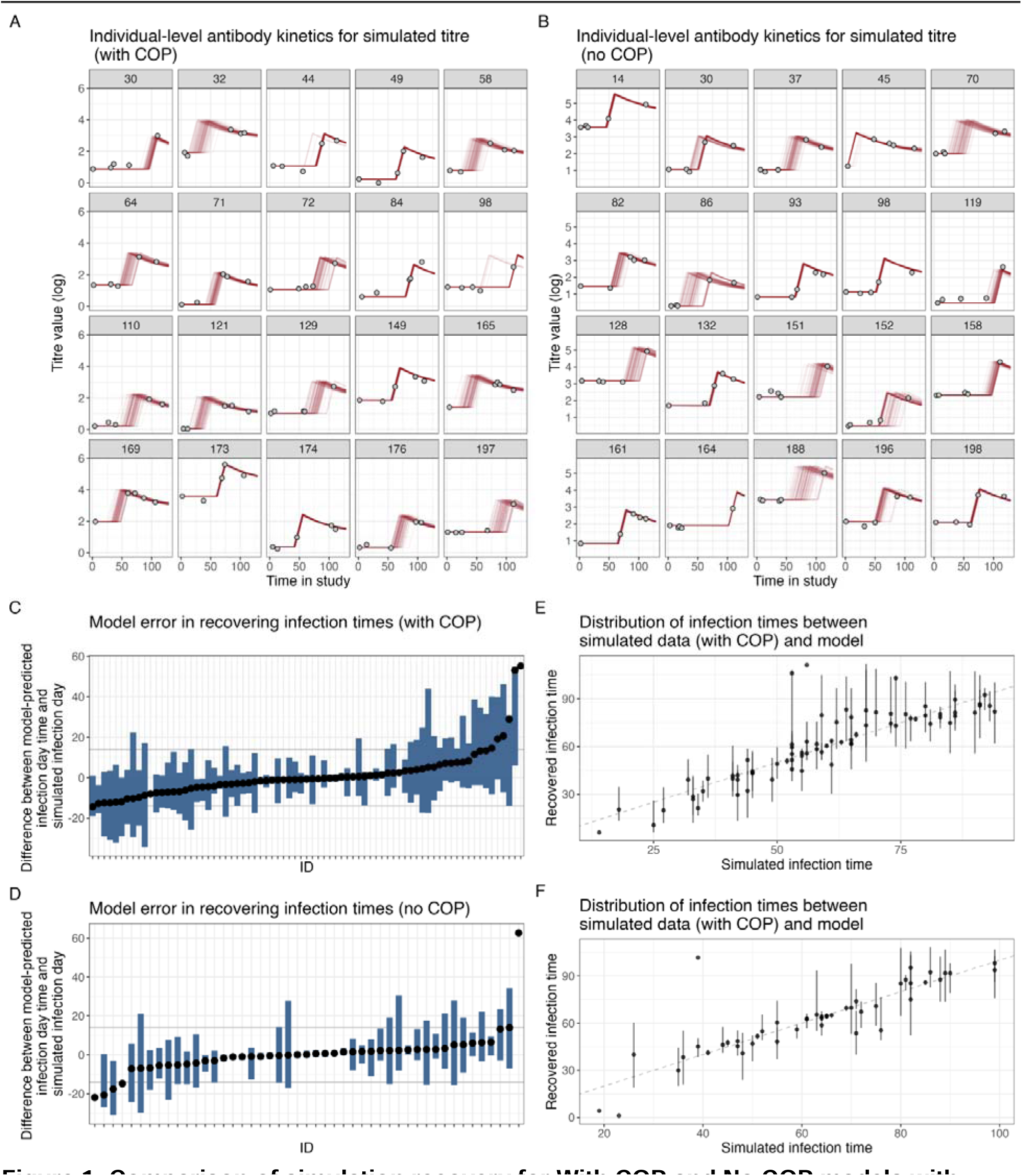
Comparison of simulation recovery for With COP and No COP models with observational error (0.1). **(A**, **B)** Individual-level antibody kinetics for a subset of individuals, showing observed data points (grey dots), and individual-level posterior medians (red lines) for With COP data and No COP data. **(C, D)** Model error in recovering infection times, represented as the difference between simulated and model-predicted infection days for infected individuals With COP data and No COP data. **(E, F)** Distribution of infection times, comparing simulated infection times (grey bars) with recovered infection times (red bars).

#### 3.1.2. Stability of simulation recovery for increasing observational noise

We assess predictive performance by measuring the Continuous Ranked Probability Score (CRPS), a metric that evaluates the accuracy of probabilistic predictions by comparing the predicted cumulative distribution function to the observed value. We find the functional form of the antibody kinetics for both No COP and With COP models is well recovered (CPRS < 0.35) across all levels of observational noise; however, decreasing in accuracy as observational noise increases (Figure 2A). In contrast, the recovery of the epidemic curve is adequate at low levels of uncertainty but quickly becomes inaccurate (CPRS > 0.7) at a moderate level of noise (Figure 2B). This suggests that for datasets with a high degree of observational error, the recovery of the epidemic timing may be inaccurate when a uniform prior on infection timing is used in *serojump*.

**Figure 2:**
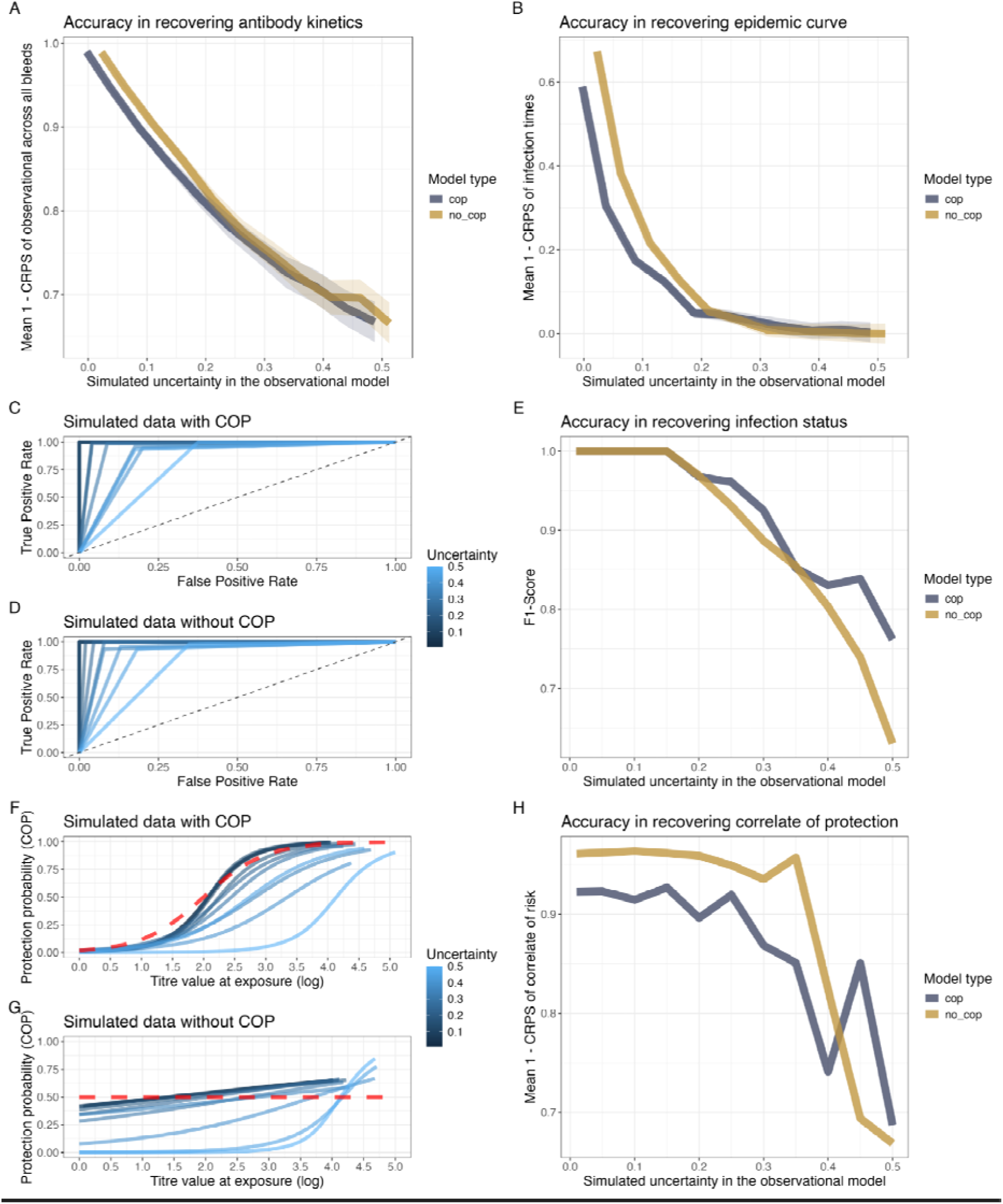
Evaluation of model performance under varying levels of observational uncertainty. **(A)** Accuracy in recovering antibody kinetics over various simulated observational errors, measured as the mean CRPS across all individuals of the observational model **(B)** Accuracy in recovering the epidemic curve over various simulated observational errors, assessed using the mean CPRS across all infection times **(C, D)** ROC curves for infection status predictions under simulated data with COP (**C**) and without COP (**D**) under different observational error (colour) **(E)** F1-scores for recovering infection status across different observational errors and model types (No COP and With COP). **(F, G)** Posterior means of the COP curves across various observational uncertainty values (blue colours) compared to the simulated function (dashed red line) **(H)** Accuracy in recovering the functional form for the COP, measured by Mean CRPS, across various levels of observational uncertainty for both model types (With COP and No NOP)

For recovery infection status, we find the model perfectly recovers the infection status of each individual (F1-score = 1) up until an observational error of 0.15, after which there is a decrease in F1-score as observational error increases, suggesting the sensitivity and specificity of the recovery of infection status is decreasing (Figure 2C–E). This decrease is driven by the increase in the false positive rate, with the true positive rate remaining consistent across all observational noise, suggesting that the model correctly identifies all infections but may incorrectly detect infections at high levels of noise. For recovery of the COP functional form, we find that when the observational noise is less than 0.3, it is well recovered for both the With COP and No COP datasets (CRPS < 0.05) (Figure 2F**–H**). However, above this observational error, the accuracy of the recovery of the functional form of the COP begins to decrease rapidly.

### 3.2 CASE STUDY 2: SEROLOGICAL INFERENCE FOR SARS-CoV-2 IN THE GAMBIA DURING THE DELTA WAVE

#### 3.2.1. Testing sensitivity of serological detection methods

We evaluate the sensitivity of various serological detection heuristics in identifying infection status. Using each of the five infection classification strategies based on serology (*serojump*, a four-fold rise in spike or NCP, and seropositivity thresholds for spike and NCP), we classified each of the PCR-confirmed individuals by their derived infection status and then calculated the sensitivity of each method (Figure 3). The ***serojump*** method demonstrates the highest sensitivity, outperforming the other four heuristics. The four-fold spike rise achieves the second-highest sensitivity, closely followed by the four-fold NCP rise. Both seropositive threshold methods (spike and NCP) perform less well, highlighting the limitations of using simple thresholds for infection detection. These observations highlight that leveraging dynamic changes in antibody kinetics can outperform static thresholds for serological detection of infection, with the ***serojump*** method demonstrating superior sensitivity, indicating its potential as a preferred approach for infection inference.

**Figure 3.**
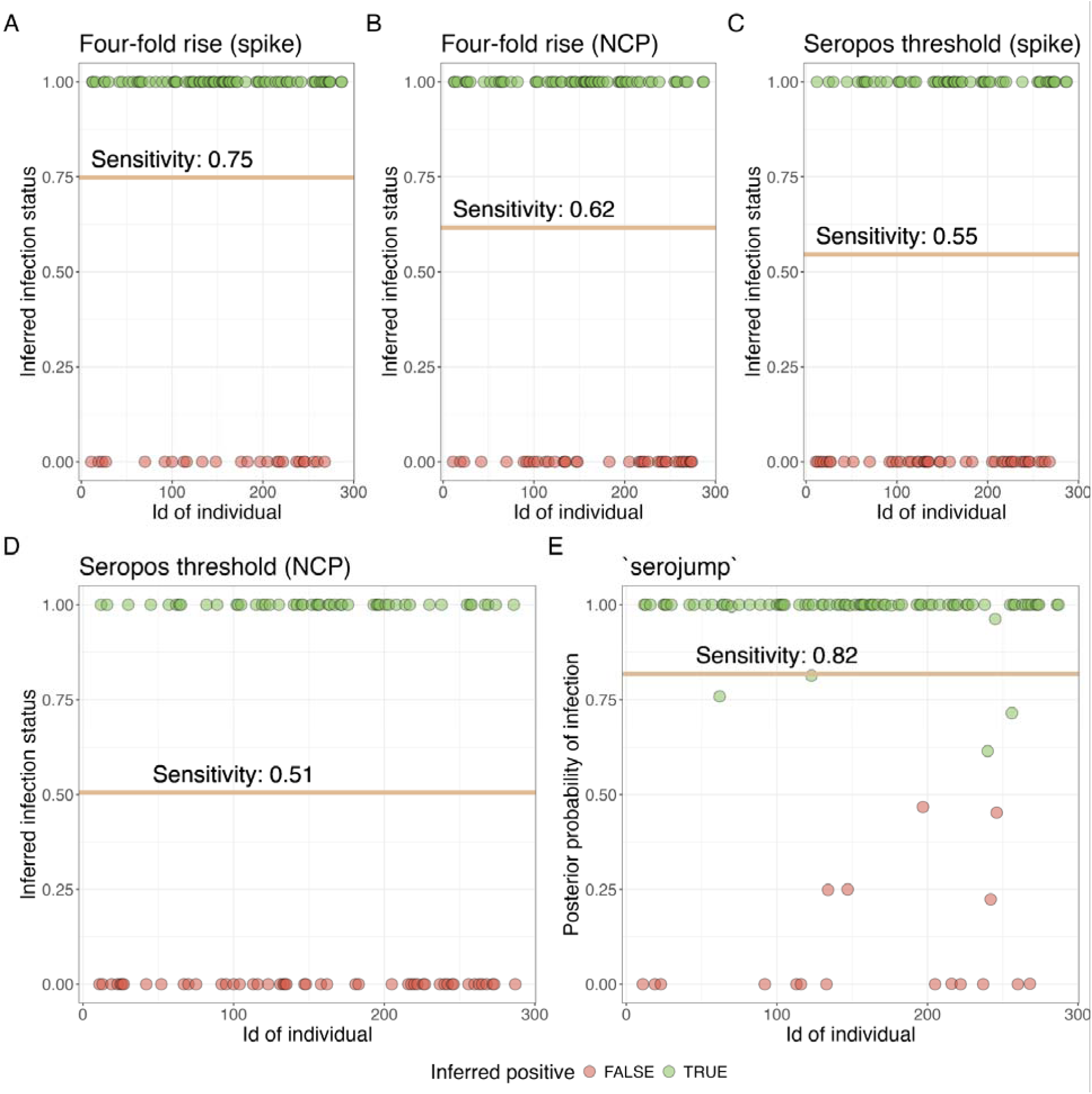
Sensitivity of serological detection heuristics in identifying infection status using serological data only. **(A)** Posterior probabilities of recovery for individuals using five detection methods: (1) Four-fold spike rise, (2) Four-fold NCP rise, (3) Seropositive threshold spike, (4) Seropositive threshold NCP, and (5) *serojump*. Green points represent individuals inferred as true positives, while red points indicate false negatives.

#### 3.2.2. Combining PCR and serological data to infer epidemiological dynamics of the Delta wave

We estimate the dynamics of antibody responses, epidemic patterns, and infection risk in the context of SARS-CoV-2 infection and vaccination during the Delta wave in The Gambia. This uses the same serological dataset described in section 3.1 but includes PCR-confirmed infections, anchoring the infection status and infection time for 99 individuals. We infer antibody kinetics trajectories for the biomarkers, spike and NCP, for individuals infected prior to the Delta variant, during the Delta wave, and following vaccination (Figure 4A). We find that infections with Delta and infections with variants before Delta stimulate both spike and NCP responses, with spike responses seeing more sustained responses compared to NCP responses (AUC: 395 for spike and 257 for NCP for Delta infection). The spike responses remain above a four-fold rise up until day 46 for infection with the Delta variant.. Following vaccination, we find robust spike responses but very low NCP responses (AUC: 607 vs 67 for spike and NCP, respectively). These observations are consistent with the spike-based composition of the vaccine used. We also summarise the total number of infections inferred from the *serojump* framework and compare this to the known PCR-confirmed infections (Figure 4B, C).

**Figure 4:**
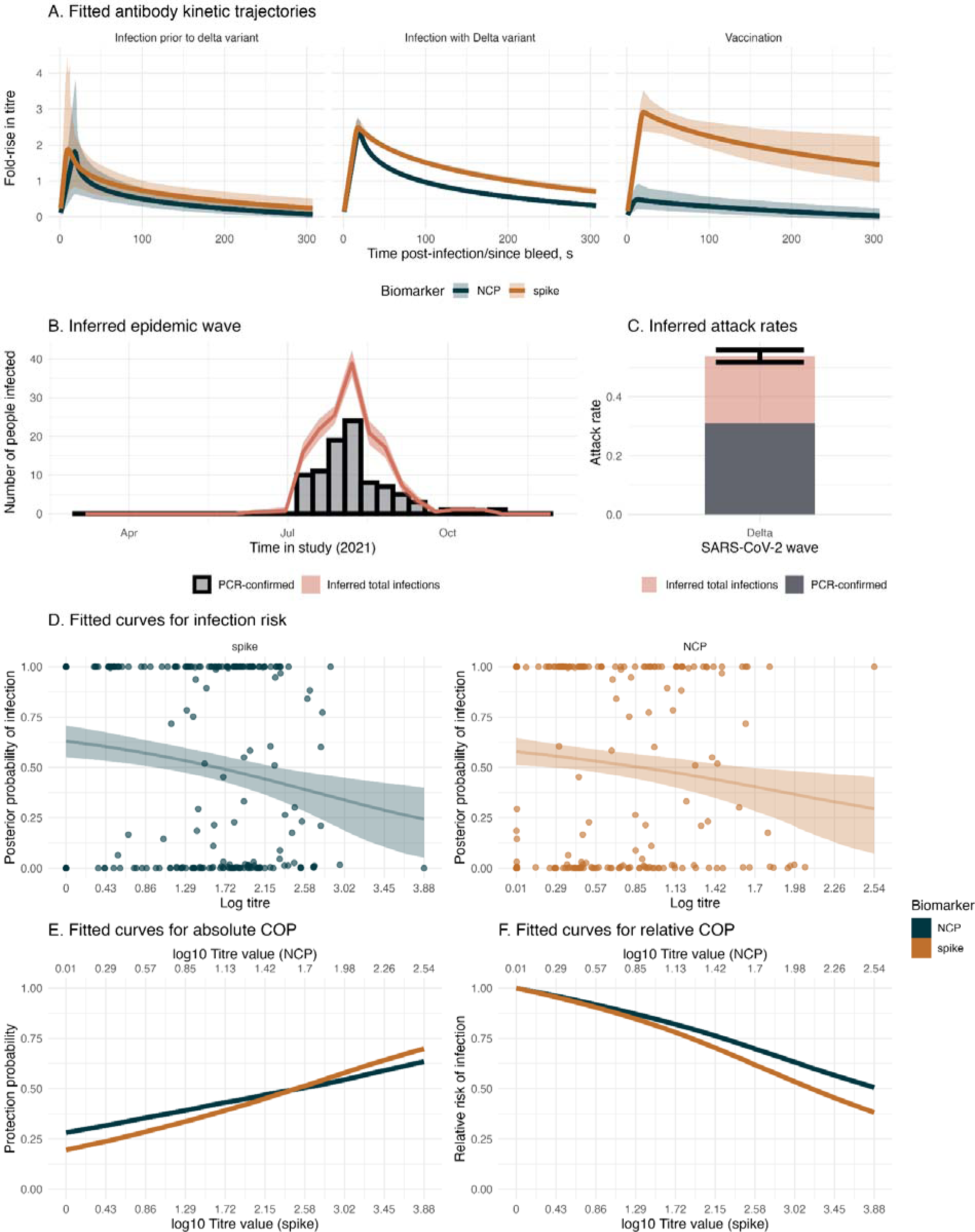
Antibody kinetics, recovery of infection timings, and correlates of protection from empirical data with PCR information. (A) Fitted antibody kinetic trajectories for individuals with infection prior to the Delta variant (left), infection during the Delta variant wave (center), and following vaccination (right), showing fold-rise in antibody titre over time for NCP (blue) and spike (orange) biomarkers. Shaded regions represent 95% credible intervals. (B) Inferred epidemic wave during the Delta wave, illustrating the temporal distribution of PCR-confirmed cases (black bars) and total inferred infections (pink shaded curve), with shaded uncertainty. (C) The estimated attack rate during the Delta wave, partitioned into PCR-confirmed cases (dark grey) and total inferred infections (pink). Error bars indicate 95% credible intervals. (D) The fitted logistic curve to the infection risk, showing the posterior probability of infection as a function of antibody titre at infection for NCP (blue) and spike (orange) biomarkers, with shaded regions representing 95% credible intervals. (E) The absolute COP from the fitted infection risk curve and (F) shows the relative COP for the same biomarkers.

Considering only PCR-confirmed infections, we find that the attack rate is 30%, increasing to 52% when we use *serojump*, implying analysing changes in serological kinetics finds many infections which are missed through virological surveillance alone. Using our informed prior ensures that the inferred timings of these missed infections agree with the known epidemic wave. Finally we compared the relationship between titre levels at infection and the individual-level posterior probability of infection, serving as a correlate of protection (COP) (Figure 4D). For NCP and spike biomarkers, a negative relationship is observed between titre levels at infection and the probability of infection, suggesting that higher antibody titres are associated with reduced infection risk. When we consider fitting the absolute COP after normalising the titre scales, spike has a slightly steeper gradient compared to NCP, suggesting a stronger COP (Figure 4E). In addition, high titre values providing only 60% protection suggest the Ancestral spike and NCP proteins correlate with protection despite being antigenically distant from the infecting variant (Delta), suggesting cross-immunity with the analysed proteins. The relative COP show that having a log spike titre of 3.2 provides 50% more protection against infection compared to those with no measurable titre (Figure 4F).

## 4. DISCUSSION

In this study, we present an open-source flexible framework for serological inference, ***serojump***, which uses a reversible-jump MCMC algorithm to probabilistically estimate whether an individual has been infected or not. Specifically, it can infer individual-level infections, antibody kinetics, and population-level dynamics using longitudinal serological data. Our approach uses mechanistic modelling of antibody kinetics to estimate latent infection states and provides an alternative to conventional serological heuristics, which are often based on crude static thresholds. Our results demonstrate that ***serojump*** can achieve high sensitivity and specificity in recovering infection status, infection timing, and population-level epidemic patterns from simulated and real-world datasets. The framework effectively captures antibody kinetics post-infection and vaccination, allowing us to differentiate between infections during different epidemic waves and post-vaccination responses. Specifically, the inferred kinetics show that vaccination induces robust spike antibody responses but negligible NCP responses, consistent with the composition of SARS-CoV-2 vaccines and previous observations.[27,28] Our results also show how static heuristics, such as four-fold rises in antibody titres or seropositivity thresholds, underperform compared to the *serojump* framework. This underscores the need for dynamic, data-driven methods to accurately infer infections, especially in scenarios where virological data are incomplete or unavailable. For example, using real-world data from The Gambia, *serojump* inferred an attack rate significantly higher than that estimated by PCR-confirmed cases alone, suggesting under-detection by virological sampling methods, potentially due to a relatively short period of peak viral shedding.[4]

Our framework addresses several challenges associated with serological studies, including variability in antibody responses and the need for flexible, biologically informed models. By integrating individual-and population-level data into a unified probabilistic framework, *serojump* offers a powerful tool for assessing pathogen transmission dynamics, evaluating vaccine efficacy, and identifying correlates of protection. These capabilities are critical for informing public health interventions, especially in resource-limited settings where virological sampling may be sparse. Furthermore, the negative relationship between antibody titres at infection and the individual-level probability of infection observed in our analysis highlights the potential for *serojump* to identify correlates of protection. These insights can guide the development of immunological benchmarks for vaccine efficacy and inform strategies for boosting population immunity. We highlight that the framework is currently pathogen agnostic and thus is adaptable to multiple pathogens with different serological profiles, highlighting the robustness of the model in handling diverse datasets, which could include varying sample sizes, intervals, and noise levels.

Inferring latent infection status and timings from serological data is a highly complex inference challenge. This requires jointly accounting for epidemic dynamics, variation in individual infection timing and status, and complex antibody kinetics that depend on the timing, nature, and antigens of infections. While it would be ideal to have a single framework capable of addressing all these mechanisms simultaneously, such an approach is challenging and demands extensive data. As a result, software tools must be tailored to address different dimensions of this complexity. *serosolver* is a general-purpose statistical inference framework that uses longitudinal and cross-sectional serological data to determine individual-level infection history, infection timings, and antibody kinetics.[19] In contrast, *serojump* focuses on estimating single infection events with precision, both in time and infection status. *serojump* also provides the ability to estimate the relationship between measured biomarkers and the probability of infection, making it particularly useful for reconstructing epidemic curves over a single epidemic. A key distinction is that *serojump* uses continuous-time sampling, and offers the user enhanced flexibility in modeling antibody kinetics and observational structures. However, *serojump* is optimized for short-term dynamics and precise infection timing rather than handling multiple infections over long timeframes or incorporating complex antigenic dynamics, which are key strengths of *serosolver*. Notably, both tools share the same underlying likelihood model,[18] but they are designed to address different types of questions in serology. Thus, these tools provide a complementary suite of methods to tackle different dimensions of serological inference challenges.

Previous studies have used Reversible-Jump MCMC methods to infer individual-level infection times and statuses from serological data.[15,16] These approaches model functional forms of antibody kinetics to estimate infection times and/or the force of infection, and subsequently establish relationships between estimated antibody titres at the time of infection and protection against disease. While these studies share similarities, such as the ability to infer multiple infections for an individual, their models and codebases are often tailored specifically to the datasets or research questions they address. As a result, they typically lack publicly available, user-friendly interfaces for broader applications to other datasets. In contrast, the methods implemented in *serojump* focus on a streamlined design that simplifies inference by only allowing a single infection to be inferred. This simplification enables a flexible and modular approach, allowing users to adapt and extend the model framework with user-written code. Unlike many existing methods, *serojump* provides a plug-and-play interface, offering general usability without requiring the re-coding of bespoke model structures. This flexibility represents a significant advance in accessibility and usability, facilitating broader adoption and application across different datasets.

Our study has limitations. The accuracy of the *serojump* framework depends on the quality of input data, including the timing, frequency, and noise levels of serological samples. However, simulation studies like the ones presented above can be used to assess the likely performance of the method with a given data structure. Our framework does not currently account for individual-level variation in kinetics (e.g., random effects) or the influence of covariates on antibody kinetics, which would be areas for future development to support research investigating these more detailed features. Scalability and runtime also pose challenges; while effective for smaller datasets, further optimization would be beneficial for applications exceeding 1,000 individuals. Potential extensions to the framework include inferring multiple infections per individual and incorporating multiple biomarkers or antigenically varied pathogens. These additions could enable the exploration of longer-term immunological phenomena (e.g., antigenic seniority)[29] and improve inference for pathogens like influenza and SARS-CoV-2. However, such extensions would significantly increase the parameter space and computational demands. Strategies such as optimizing samplers or integrating population-level MCMC algorithms (e.g., parallel tempering) could mitigate these challenges and allow for more complex hierarchical frameworks to be evaluated.

The development and implementation of *serojump* address key limitations of previous methods used to infer infection times and statuses from serological data. While Reversible-Jump MCMC and related approaches have long been established for modeling latent infection states, their application has often been constrained by bespoke and complex implementations that lack generalizability. By contrast, *serojump* introduces a user-friendly, modular framework that simplifies inference to a single infection event while providing the flexibility for users to customize and extend the model for diverse datasets and research questions. This approach not only democratises the use of advanced serological modeling techniques but also lays the foundation for broader applications and collaborative development in the field. The ability to balance simplicity with flexibility ensures that *serojump* is well-positioned to support future advancements in understanding infection dynamics and serological data interpretation.

Therefore, we developed a general-purpose statistical framework that leverages reversible jump MCMC to systematically infer antibody kinetics, infections and their timing using individual-level antibody kinetic for an epidemic outbreak. This tool provides a robust, flexible approach to better inform epidemiological parameters from serological data. By making these advanced methods more accessible and adaptable in the form of an R package *serojump*, we hope this framework will substantially enhance our ability to track and control infectious diseases in diverse settings.

## Financial Disclose Statement

The study was funded by a United Kingdom Research and Innovation Grant (No. MC_PC_19084). JAH is supported by a Wellcome Trust Early Career Award (grant 225001/Z/22/Z). AJK is supported by National Institutes of Health (1R01AI141534-01A1) and Wellcome (226142/Z/22/Z).

## Data Availability

All data produced are available online at https://seroanalytics.org/serojump/

## SUPPLEMENTARY FIGURE CAPTIONS

### Supplementary Methods. Supporting documentation for them methods section

**Figure S1: Convergence diagnostics for simulated data with COP and 0.1 uncertainty in observational error. (A)** Trace plots for fitted parameters (sigma, wane, a, b, c) across four Markov chains, illustrating the mixing and convergence of the parameters. **(B)** Trace plots for the log posterior across the four chains, showing the variability and stabilization of the log posterior over iterations. **(C)** Convergence diagnostics for fitted parameters, including effective sample size (ess_bulk, ess_tail) and R^hat^, which assess the adequacy of sampling and convergence for each parameter. **(D)** Trace plots for transdimensional convergence of the model dimension, with histogram counts of model dimensions sampled across the chains. **(E)** Trace plots for transdimensional convergence for the SMI (Structural Model Index) and histogram counts of the log-transformed SMI values across chains. **(F)** Convergence diagnostics for transdimensional parameters, including effective sample size (ess_bulk, ess_tail) and R^hat^, summarizing the adequacy of sampling and convergence for the transdimensional space.

**Figure S2: Convergence diagnostics of infection timing for simulated data with COP and 0.1 uncertainty in observational error. (A)** Trace plots for the timing of infection for individuals with posterior P(Z) > 0.5 display estimates across four Markov chains. Each point and its uncertainty interval reflect the sampled infection timing for each individual over iterations. **(B)** Convergence diagnostics for the timing of infection for individuals with posterior P(Z)>0.5, showing R^hat^ values for each individual. The red dashed line indicates the threshold for R^hat^=1.1, which marks convergence.

**Figure S3: Convergence diagnostics for simulated data No COP and 0.1 uncertainty in observational error. (A)** Trace plots for fitted parameters (sigma, wane, a, b, c) across four Markov chains, illustrating the mixing and convergence of the parameters. **(B)** Trace plots for the log posterior across the four chains, showing the variability and stabilization of the log posterior over iterations. **(C)** Convergence diagnostics for fitted parameters, including effective sample size (ess_bulk, ess_tail) and R^hat^, which assess the adequacy of sampling and convergence for each parameter. **(D)** Trace plots for transdimensional convergence of the model dimension, with histogram counts of model dimensions sampled across the chains. **(E)** Trace plots for transdimensional convergence for the SMI (Structural Model Index) and histogram counts of the log-transformed SMI values across chains. **(F)** Convergence diagnostics for transdimensional parameters, including effective sample size (ess_bulk, ess_tail) and R^hat^, summarizing the adequacy of sampling and convergence for the transdimensional space.

**Figure S4: Convergence diagnostics of infection timing for simulated data No COP and 0.1 uncertainty in observational error. (A)** Trace plots for the timing of infection for individuals with posterior P(Z) > 0.5 display estimates across four Markov chains. Each point and its uncertainty interval reflect the sampled infection timing for each individual over iterations. **(B)** Convergence diagnostics for the timing of infection for individuals with posterior P(Z)>0.5, showing R^hat^ values for each individual. The red dashed line indicates the threshold for R^hat^=1.1, which marks convergence.

**Figure S5: Convergence diagnostics for empirical data without PCR information (A)** Trace plots for fitted parameters (sigma, wane, a, b, c) across four Markov chains, illustrating the mixing and convergence of the parameters. **(B)** Trace plots for the log posterior across the four chains, showing the variability and stabilization of the log posterior over iterations. **(C)** Convergence diagnostics for fitted parameters, including effective sample size (ess_bulk, ess_tail) and R^hat^, which assess the adequacy of sampling and convergence for each parameter. **(D)** Trace plots for transdimensional convergence of the model dimension, with histogram counts of model dimensions sampled across the chains. **(E)** Trace plots for transdimensional convergence for the SMI (Structural Model Index) and histogram counts of the log-transformed SMI values across chains. **(F)** Convergence diagnostics for transdimensional parameters, including effective sample size (ess_bulk, ess_tail) and R^hat^, summarizing the adequacy of sampling and convergence for the transdimensional space.

**Figure S6: Convergence diagnostics for empirical data without PCR information. (A)** Trace plots for the timing of infection for individuals with posterior P(Z) > 0.5 display estimates across four Markov chains. Each point and its uncertainty interval reflect the sampled infection timing for each individual over iterations. **(B)** Convergence diagnostics for the timing of infection for individuals with posterior P(Z)>0.5, showing R^hat^ values for each individual. The red dashed line indicates the threshold for R^hat^=1.1, which marks convergence.

**Figure S7: Convergence diagnostics for empirical data with PCR information (A)** Trace plots for fitted parameters (sigma, wane, a, b, c) across four Markov chains, illustrating the mixing and convergence of the parameters. **(B)** Trace plots for the log posterior across the four chains, showing the variability and stabilization of the log posterior over iterations. **(C)** Convergence diagnostics for fitted parameters, including effective sample size (ess_bulk, ess_tail) and R^hat^, which assess the adequacy of sampling and convergence for each parameter. **(D)** Trace plots for transdimensional convergence of the model dimension, with histogram counts of model dimensions sampled across the chains. **(E)** Trace plots for transdimensional convergence for the SMI (Structural Model Index) and histogram counts of the log-transformed SMI values across chains. **(F)** Convergence diagnostics for transdimensional parameters, including effective sample size (ess_bulk, ess_tail) and R^hat^, summarizing the adequacy of sampling and convergence for the transdimensional space.

**Figure S8 Convergence diagnostics for empirical data with PCR. (A)** Trace plots for the timing of infection for individuals with posterior P(Z) > 0.5 display estimates across four Markov chains. Each point and its uncertainty interval reflect the sampled infection timing for each individual over iterations. **(B)** Convergence diagnostics for the timing of infection for individuals with posterior P(Z)>0.5, showing R^hat^ values for each individual. The red dashed line indicates the threshold for R^hat^=1.1, which marks convergence.

**Figure S9: Antibody kinetics, recovery of infection timings, and correlates of protection from empirical data without PCR information. (A)** Fitted antibody kinetic trajectories for individuals with infection prior to the Delta variant (left), infection during the Delta variant wave (center), and following vaccination (right), showing fold-rise in antibody titre over time for NCP (blue) and spike (orange) biomarkers. Shaded regions represent 95% credible intervals. **(B)** Inferred epidemic wave during the Delta wave, illustrating the temporal distribution of PCR-confirmed cases (black bars) and total inferred infections (pink shaded curve), with shaded uncertainty. **(C)** The estimated attack rate during the Delta wave, partitioned into PCR-confirmed cases (dark grey) and total inferred infections (pink). Error bars indicate 95% credible intervals. **(D)** The fitted logistic curve to the infection risk, showing the posterior probability of infection as a function of antibody titre at infection for NCP (blue) and spike (orange) biomarkers, with shaded regions representing 95% credible intervals. **(E)** The absolute COP from the fitted infection risk curve and **(F)** shows the relative COP for the same biomarkers.

