## Supplementary Methods for "*serojump*: A Bayesian tool for inferring infection timing and antibody kinetics from longitudinal serological data"

1. **Reversible-Jump Markov Chain Monte Carlo (RJ-MCMC) Algorithm on Serological Data**

1.1. Sampling from the Posterior Distribution

1.2. Proposal Distributions

1.3. Serojump Algorithm
1.4. Choice of Priors
1.5. Modifications for Known Infections
1.6. Postprocessing of the Posterior Distributions
1.7. Infection risks and correlates of protection

1.8 Assessing convergence of the posterior distribution

2. **The Model Specification in Serojump for Simulated Data**
   2.1. Data Overview
   2.2. Model Overview
   2.3. Implementation
3. **The Model Specification in Serojump for Empirical Data**
   3.1. Data Overview
   3.2. Model Overview (A: No Known Infections, B: Some Known Infections)
   3.3. Implementation

SUPPLEMENTARY METHODS

1. Reversible-Jump Markov chain Monte Carlo (RJ-MCMC) algorithm on serological data

1.1 SAMPLING FROM THE POSTERIOR DISTRIBUTION

To sample from EQUATION 2 using a Metropolis-Hasting algorithm, we must define proposal distributions for $(\tau,Z,\theta)$. However, $\tau$ does not have a fixed length throughout this sampling procedure as its length depends on the number of individuals who are infected, the vector describing which (Z) is also sampled and thus changes dynamically through the algorithm. Therefore, it is impossible to use the Metropolis-Hasting algorithm, as the posterior distribution does not have a fixed number of dimensions, violating the conditions of the Ergodic Theorem.

The RJ-MCMC algorithm allows sampling across transdimensional posterior distribution by adjusting the Metropolis-Hasting acceptance ratio when the dimension of a proposed state varies from the current state(Green 1995). In the context of our posterior distribution (EQUATION 2), when the proposed number of infections differs from the current number of infections, it allows the model to sample across all these spaces whilst ensuring the condition of the Ergodic Theorem is not violated. Though it is technically possible to jump from any current state $(\tau,Z,\theta)$ to any proposed state $(\tau',Z',\theta')$, for mathematical simplicity, we consider three proposal potential regimes which closely relate to the proposed and current state. These three regimes are a birth step, a death step, and a neutral step.

1.1.1 *Birth step*

In a birth step, the proposed state includes an additional infected person compared to the current state, this is achieved by sampling a person, *j*, uniformly from the currently uninfected individuals; **j** $\sim$ Uniform**(Z^0^),** where **Z^0^ = {i | Z_i_ = 0},** and updating Z $\to$ Z’. Then sampling an infection timing from $\tau_{j}'$~ P_exp_, and updating $\tau' = \tau\cup\tau_{j}',$and sampling priors from a proposal $\theta'$~ q_1_($\theta$). Then, given the proposed state (Z’, $\tau$’, $\theta$’) the acceptance ratio of this new infection is given by (derivation in Appendix)

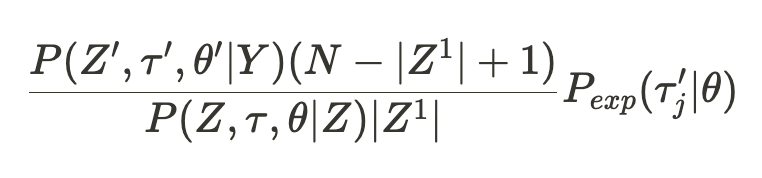

EQUATION 2. Acceptance ratio of the RJ-MCMC for a birth step.

Where Z**^1^ = {i | Z_i_ = 1} and |.|** is the magnitude of the vector.

1.1.2 *Death step*

In a death step, the proposed state includes one less infected individual compared to the current state and is achieved by sampling a person, *j*, uniformly from the currently infected individuals;

**j** $\sim$ **Uniform(Z^1^), where Z^1^ = {i | Z_i_ = 0},**

and updating Z $\to$ Z’ and the vector $\tau$ accordingly and sampling from the proposal $\theta'$~ q_1_($\theta$), the proposed state $(\tau',Z',\theta')$ the acceptance ratio of this new infection is given by

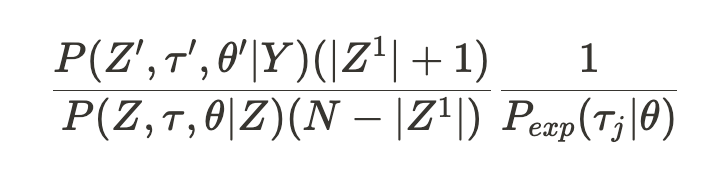

EQUATION 3. Acceptance ratio of the RJ-MCMC for a death step.

1.1.3 *Neutral step*

In a neutral step, the infection status of individuals is unchanged, but we randomly sample an infection time from a proposal distribution, ${\tau'}_{j}\sim$q_2_($\tau$_j_) for individual j $\sim$ **Uniform(Z^1^),** and $\theta'$~ q_1_($\theta$). Therefore, the acceptance ratio is given by the usual Metropolis-Hastings algorithm.

1.2. PROPOSAL DISTRIBUTIONS q_1_($\theta$), q_2_($\tau$_j_)

1.2.1 Fixed variables, q_1_($\theta$)

We use an adaptive proposal distribution q_1_($\theta$) to sample the parameter space θ. The adaptive Metropolis-Hastings algorithm provides a systematic method for modifying the shape of the proposal distribution based on the accepted steps of the current Markov chain, allowing for more efficient mixing of chains. That is, the q_1_(θ^(i)^) = N(θ^(i)^,Σ^(i)^(θ^(i)^)) follows a Gaussian distribution with the covariance matrix Σ^(i)^. The Markov chain runs for an initial number of steps (T_init_) from a truncated multivariate normal proposal distribution with a covariance matrix, I_s_, whose entries are calculated using the upper and lower bounds of the support of the priors [s_k_^0^, s_k_^1^] ∈ S, through i_k,k_=(s_k_^1^−s_k_^0^)/ζ and i_i,j_=0 otherwise, where ζ is a scaling factor.

Problematically, the proposal distribution using the updated covariance matrix, Σ^(i)^, is no longer memoryless, and therefore, the chain may no longer converge to the correct stationary distribution. To overcome this problem, the proposal distribution must also sample from a non-adaptive multivariate Gaussian distribution modified to ensure that changes to the covariance matrix diminish over time. Further, to improve chain mixing and to optimise convergence rates, we include adaptive scaling factors, λ(i) and M(i), for the initial non-adaptive and adaptive proposals, respectively, whose magnitude diminishes with the number of steps in the chain. The adaptive scaling factor for the non-adaptive proposal distributions stops once the model starts sampling from the adaptive proposal distributions. Overall, the combined non-adaptive and adaptive proposal distributions for the adaptive Metropolis-Hastings is given by:

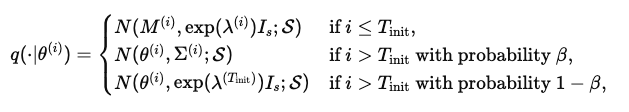

Where Σ^(i)^ = exp(M^(i)^)Γ^(i)^ and M^(i)^, λ^(i)^, and Γ^(i)^ are updated iteratively through the stochastic approximation algorithm:

λ^(i+1)^=λ^(i)^+γ_1_(i)(α(θ^(i)^,θ′)−0.234),

M^(i+1)^=M^(i)^+γ_2_(i)(α(θ^(i)^,θ′)−0.234),

μ^(i+1)^=μ^(i)^+γ_3_(i)(μ^(i)^−θ^(i)^),

Γ^(i+1)^=Γ^(i)^+γ_4_(i)(θ^(i)^−μ^(i+1)^)(θ^(i)^−μ^(i+1)^)^T^−Γ^(i)^

Here γ_x_(i) are gain factors. Note that when i>T_init_, we stop updating λ(i).

In our implementation, we define $\psi_{adapt}^{(i)}=\{M^{(i)},\mu^{(i)},\Gamma^{(i)},\lambda^{(i)} \}$ and choose values β=0.95, ζ=100, λ^(0)^=log⁡(0.12/∣θ∣), M^(0)^=log⁡(2.382^2^/∣θ∣), μ(0)=π_0_, I_s_=I (identity matrix),and γ_x_(i)=(1+i)^−0.5^ for all x.

1.2.2. Timing of infection, q_2_($\tau$_j_)

To allow efficient sampling for $\tau_{j}$ in the neutral step, we defined a proposal distribution which is adaptively updated but centred around the current value, $\tau$’_j_ $\sim$q_2_($\tau$_j_) = N($\tau$_j_, σ_i_) where σ_i_ is updated according to the algorithm;

log(σ_i_) $\leftarrow$ log(σ_i_) + (1 + k)^-0.5^($\alpha$ – 0.44)

Where *k* is the interaction in the Markov chain and $\alpha$ is the acceptance ratio for that proposal. Initially we choose σ_i_^(0)^ = 1.

1.3. *serojump* ALGORITHM

With these regimes and the proposal distributions defined, we can define the algorithm for sampling from the posterior.

**Algorithm 4.1: Birth-Death Reversible Jump MCMC Algorithm**

1. Choose a model *k* and initialise the chain with an initial state {θ^(0)^, τ^(0)^, Z^(0)^}
2. For i = 1 to N, do:
   - Determine p_birth_, p_death_, p_par_
     i. If 0< |τ^(0)^| <M, then p_birth_=p_death_=p_par_=0.33
     ii. If |τ^(0)^| = 0, then p_birth_=0.67, p_death_=0, p_par_=0.33,
     iii. If |τ^(0)^| = M, then p_birth_=0, p_death_=0.67, p_par_=0.33,
   - Sample $\theta$’ ~ q_1_($\theta$^(i)^| $\psi_{adapt}^{(i)}$), u_1_ ~ Uniform(0, 1)
   - If u_1_ < p_birth_, then
     i. Select j′∼Uniform({j ∣ Z_j_^(i)^=0}) and set Z_j’_’ = 1, τ_j_’ ~ P_exp_(τ^(i)^) and update {θ^(i)^, τ^(i)^, Z^(i)^} -> {θ^(1)^, τ’, Z’}
     ii. Calculate the acceptance probability, α, given by Equation 1.1
   - Else if u_1_ < (p_birth +_ p_death_), then
     i. Select j′∼Uniform({j ∣ Z_j_^(i)^=1}) and set Z_j’_^’^ = 0.
     ii. Calculate the acceptance probability, α, given by Equation 1.2
   - Else
     i. Select j′∼Uniform({j ∣ Z_j_^(i)^=1}), resample τ_j’_’ ~ P_exp_(τ^(i)^, σ_j’_^(i)^) and update {θ^(i)^, τ^(i)^, Z^(i)^} -> {θ^(1)^, τ’, Z’}
     ii. Compute acceptance probability, α, given by Equation 1.3
   - Update adaptive proposal distributions σ_j’_^(i+1)^ <- σ_j’_^(i)^, $\psi_{adapt}^{(i+1)}\leftarrow\psi_{adapt}^{(i)}$
   - Sample u_2_ ~ Uniform(0, 1)
   - If u_2_ < α:
     i. Acceptance candidate state {θ^(i+1)^, τ^(i+1)^, Z^(i+1)^} <- {θ’, τ’, Z’^)^}
   - Else
     i. Reject candidate state {θ^(i+1)^, τ^(i+1)^, Z^(i+1)^} <- {θ^(i)^, τ^(i)^, Z^(i)^}
3. End For.

Using the algorithm defined, we devise an algorithm that permits efficient sampling across the transdimensional posterior distribution P($\tau,Z,\theta$ | Y).

1.4. CHOICE OF PRIORS ON P(Z), P_exp_($\tau$_i_), P($\theta$)

The user is required to input a distribution for P_exp_($\tau$_i_), if not given, it will assume a uniform distribution across the study period [P_exp_($\tau$_i_) ~ Uniform(1, T)] for each individual. For P(Z), the trans-dimensional sampling procedure induces a strong implicit prior on the number of infected persons of |Z^1^| ~ Bin(N, 0.5). To remove this prior and force every individual to have a uniform chance of infection, we negate this effect by choosing a prior on P(Z) = $\left( N \right)$(N |Z^1^|)^-1^. This effectively introduces an individual-level probability of infection of U(0, 1) for each individual. We can make this prior informative, say if we think each individual has a probability of infection of *p*, then we choose a prior of P(Z) = (N |Z^1^|)^-1^ x Bin(Z | N, p). The priors on the parameters driving the antibody kinetics and observational model P($\theta$), are defined by the user.

1.5. MODIFICATION FOR KNOWN INFECTIONS

When known infections are included in the model, and the model is used to infer missed infections, the algorithm is modified in several ways. First, by defining K as the vector of individuals for which infection is known and |K| the number of known infections, then the sampling procedure over I and $\tau$ becomes constrained to those not infected K’. Consequently, this changes the N -> N-K in the transdimensional convergence steps, and the implicit prior on |Z^1^| ~ Bin(N - K, 0.5). These changes are dealt with automatically within the *serojump* package.

1.6 POSTPROCESSING OF THE POSTERIOR DISTRIBUTIONS

After fitting the *serojump* algorithm, we have posterior distributions for the space $(\hat{\tau}, \hat{Z}, \hat{\theta})$. The posterior distributions $\hat{\theta}$ are fixed in dimensions and continuous values and are assessed and plotted for convergence as usual. For $\hat{Z}$, our posterior distribution is a binary vector of values representing infected and not-infected of a fixed length. And finally $\hat{\tau}$, whose values are continuous but posterior distributions are not of a fixed length and correspond to the infections in $Z$. To assess the posterior distribution of the post-exposure antibody trajectories for exposure *e*, and biomarker *b*, we calculate $f_{e}^{b}(t, \hat{\theta})$. To assess the posterior distribution of the number of infected individuals across the population, we calculate $\sum_{i=1}^{N} \hat{Z_{i}}$ for each sample of the posterior distribution. The posterior distributions of the timings of infection are calculated from $\hat{\tau_{i}}$ for each individual, noting that the size of the $\hat{\tau_{i}}$ may vary between 0 and M, depending on how often this individual is determined to be infected in the posterior distributions $\hat{Z}$.

1.7 INFECTION RISKS AND CORRELATES OF PROTECTION

1.7.1. REFERENCE TITRE VALUES

For each posterior sample, we first calculate “reference titre value” for each individual, which for those infected $\hat{Z_{i}}$ = 1 is the titre at which individuals become infected (time τ) for each individual s_i_ = (A_i, τ_^b^). For those not infected, we sample a random value from their titre trajectories across the study weighted by the prior probability of exposure timing w($\tau$) = (P_exp_($\tau$_i_). Once calculated for all the posterior samples, for each individual, we calculate the expectation of both the infection status p_i_ = E[ $\hat{Z_{i}}$ ] and for the reference titre value E[s_i_].

1.7.2. DETERMINING THE COP WHERE EXPOSURE IS KNOWN (SIMULATED DATA)

In the simulated data, we have the exposure rate simulated. To check the accuracy of the algorithm at the recovery of the simulated COP, we fit a logistic curve of the form:

$$R(x) = L(1-\frac{1}{1+exp(-k(x - x_{0}))})$$

To the infection risk. Here the response variable is the expectation of the infection status (p_i_), and the explanatory variable (x) is the expectation reference titre value for each individual E[s_i_], and where L is the exposure rate, which is simulated as 0.65 for the COP model and 0.55 for the No COP. The COP curve is then given by $aCOP(x) = \frac{1}{1+exp(-k(x - x_{0}))}$.

The fitting procedure is done in *stan* with the prior distributions, k ~ *N(0, 5),* x_0_ ~ *N(mid(x), mid(x)/s)* and assuming a normal likelihood with a fitted standard deviation sigma ~ *N^+^(0, 1)*.

1.7.3. DETERMINING THE COP WHERE THE EXPOSURE STATUS IS UNKNOWN (EMPIRICAL DATA)

To determine the relative correlate of protection (rCOP) and absolute correlate of protection (aCOP) from the serojump algorithm, we use the reference titre values and fit a logistic curve above but instead of fixing L, we also fit this value too, with the prior L ~U(0, 1). Once fitted, we can extract the absolute correlate of the protection curve as above, $aCOP(x) = \frac{1}{1+exp(-k(x - x_{0}))}$. The relative COP is calculated by finding the value of R(X = min(x)) and then finding f[min(x)], and then calculating

.

$$rCOP(x) = \frac{R(x)}{R[min(x)]}$$

1.8 ASSESSING CONVERGENCE OF THE POSTERIOR DISTRIBUTIONS

To assess the convergence of our fitted posterior space $(\hat{\tau}, \hat{Z}, \hat{\theta})$, we can simply use the usual Gelman-Rubin statistics for $\hat{\theta}$, which is a fixed dimensional space of continuous vectors. To show that ($\hat{\tau}, \hat{Z})$ we have converged, we use a transdimensional convergence statistic which considered convergence of $\sum_{i=1}^{N} \hat{Z_{i}}$ and the convergence of $\hat{\tau}$ through an scale model indicator (SMI) algorithm.(Somogyvári and Reich 2020) Showing that both $\sum_{i=1}^{N} \hat{Z_{i}}$ and SMI of $\hat{\tau}$ have converged is sufficient to determine the convergence of the *serojump* algorithm.

2. The model specification in *serojump* for simulated data

2.1 DATA OVERVIEW

| **Variable** | **Values** |
| --- | --- |
| Number of individuals (i) | {1, 2, …, 200} |
| Length of study period (t) | {1, 2, …, 120} |
| Biomarkers measured (b) | IgG |
| Exposure types considered | Pre-infection ($e_{pre})$  Post-infection ($e_{inf})$ |
| Fitted exposure type | Infection (with no known) |

Table SM1. Overview of the serological data in the simulated dataset.

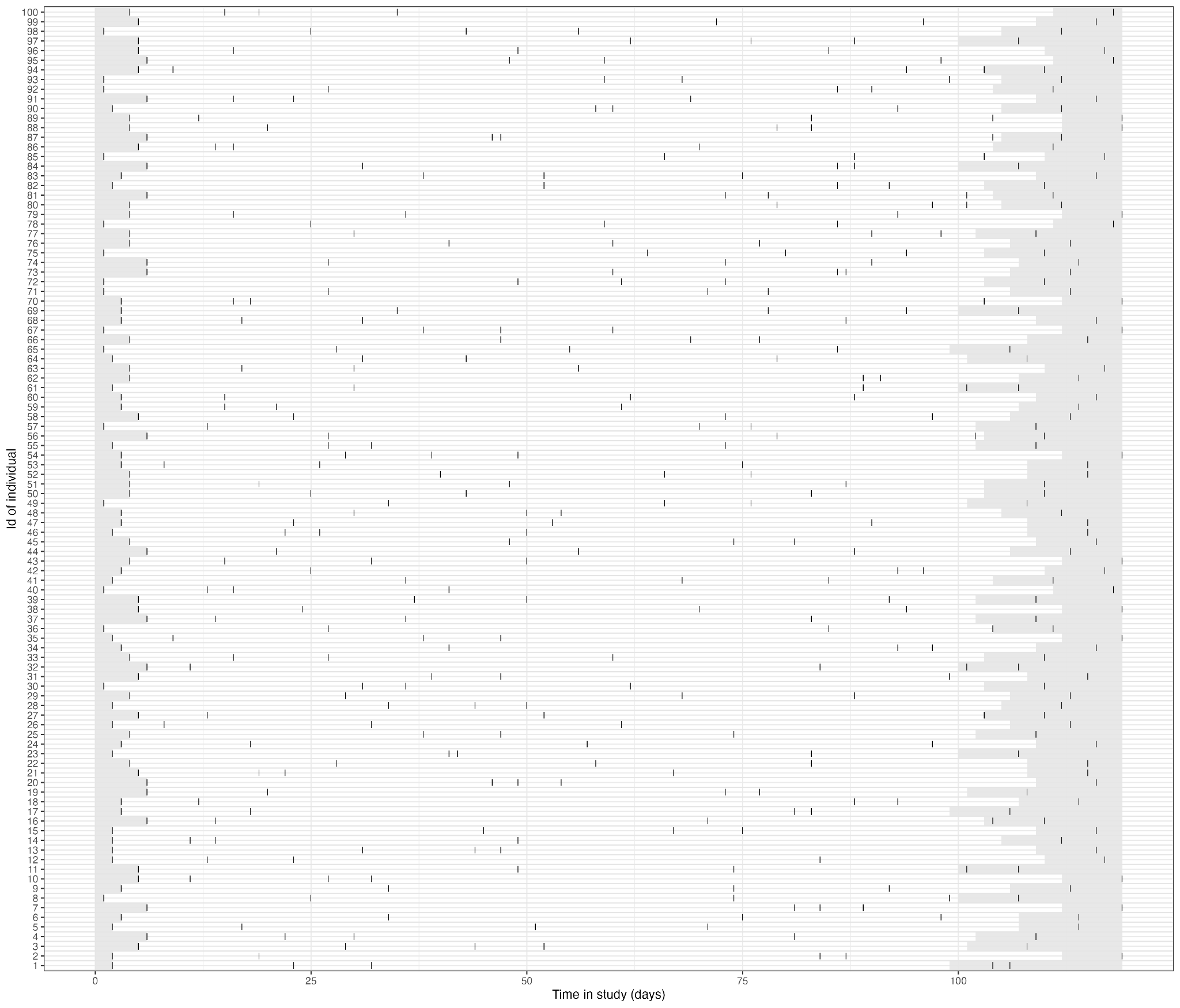

Figure SM1. **Longitudinal timeline of individual follow-up in the study.** Each row represents a unique individual (showing first 100), with the x-axis showing the time in the study (days). Gray bars indicate the period where an infection cannot be inferred, while black points mark dates that serological samples were taken.

2.2. MODEL OVERVIEW

| OBSERVATIONAL MODEL: $P_{obs}(Y_{i, t}^{b}\vert A_{i,t}^{b},\theta)$ | | |
| --- | --- | --- |
|  | Likelihood function: | P_obs_ = *f_N_*(A_i, t_^b^\| Y_i, t_^b^, 𝜎)  Priors: 𝜎 ~ Uniform(0, 4) |
| ANTIBODY KINETICS: $f_{e_{pre}}$,$f_{e_{inf}}$, (see Figure for PP) | | |
|  | Pre-infection (e_pre_) | **Functional** Form:$f_{e_{pre}}(t, Y_{i,0}) =Y_{i,0} - tw$  **Priors**: w ~ Uniform(0, 0.1) |
|  | Post-infection (e_inf_) | **Functional Form:**  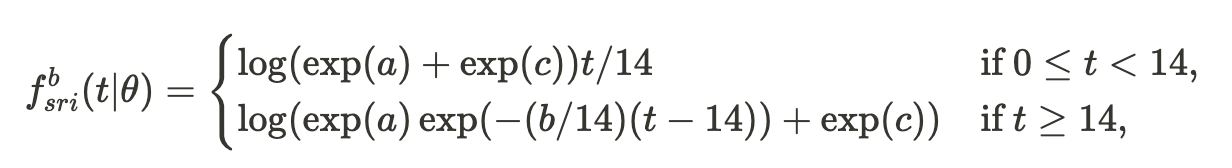  **Priors**: a ~ Normal(2, 2), b ~Normal(0.3, 0.05), c ~ Normal(0, 4). |
| PRIOR ON INFECTION TIME, $P_{exp}(\tau_{i})$ | | |
|  | For each individual, *i* | $P_{exp}(\tau_{i}) \sim Uniform(1, 120)$ |
| PRIOR ON TOTAL NUMBER OF INFECTIONS, $P(Z)$ | | |
|  | For each individual , *i* | Z_i_ ~ Bernoulli(0.5) |

Table SM2. Overview of the model inputs for *serojump* for the simulated dataset.

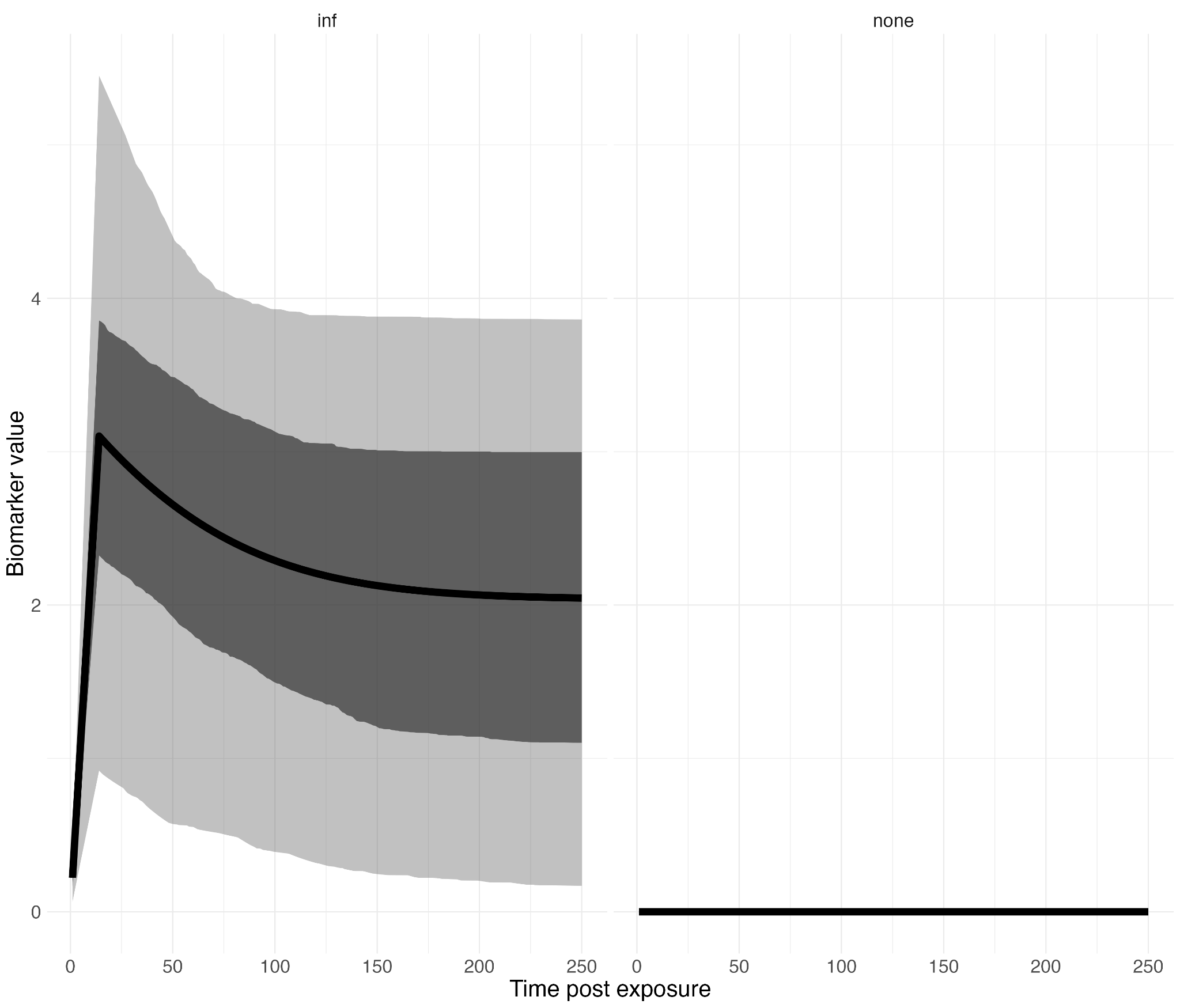

**Figure SM2. Prior predictive distributions for antibody kinetics.** The solid black line represents the median predicted antibody response over time, while the shaded area shows the uncertainty (e.g., 95% credible interval) in the prior predictions. The x-axis represents time (days), and the y-axis indicates the modeled antibody level. We show the distributions for infection and pre-infection.

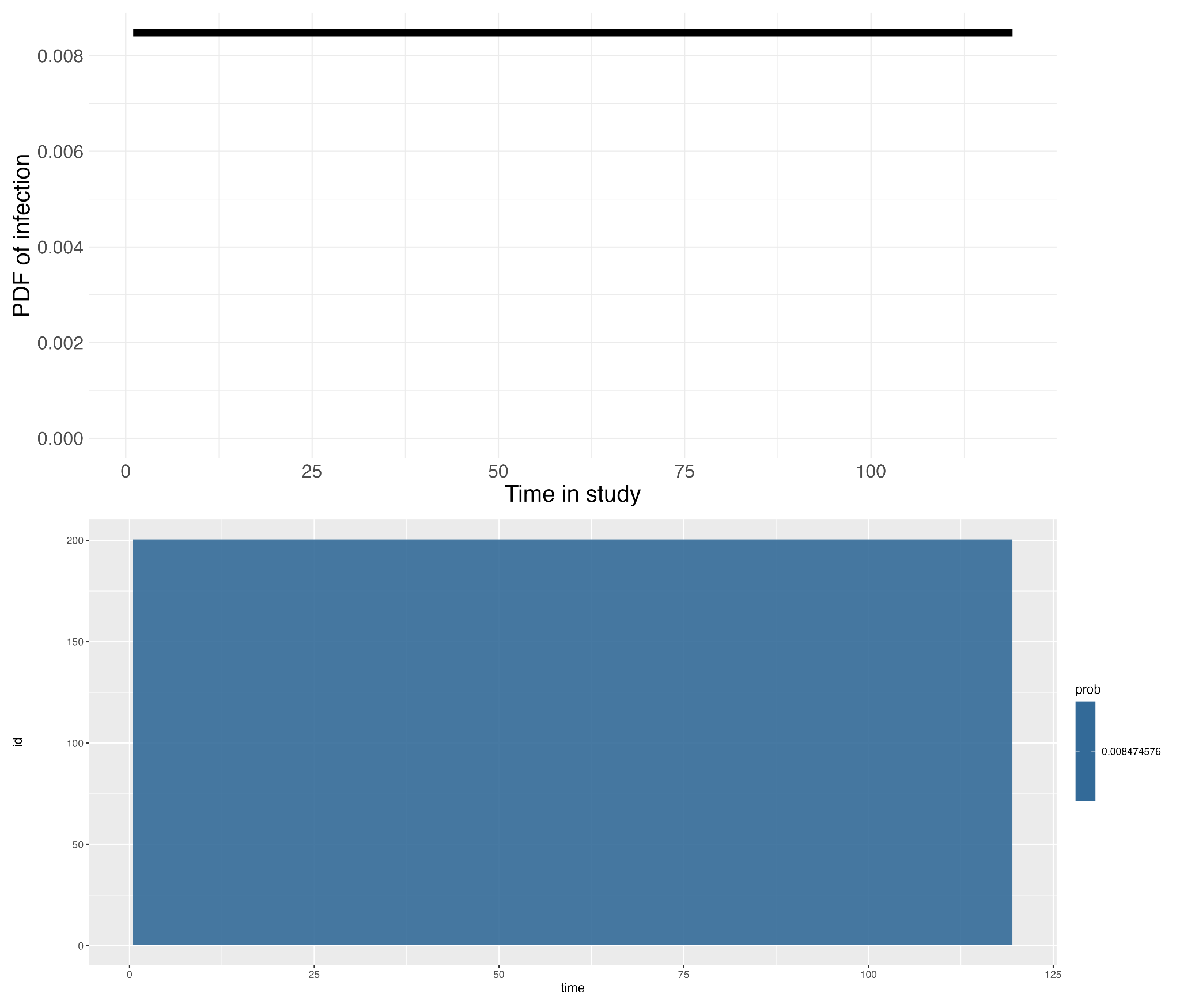
**Figure SM3. Probability density and individual-level posterior probabilities of infection over time.** The top panel shows the probability density function (PDF) of infection events across the study period (empirical probability density function). The x-axis represents time in the study (days), and the y-axis represents the PDF of infection. The bottom panel provides a heatmap of posterior probabilities of infection for each individual (y-axis) over time (x-axis). In this case, there is a uniform probability of infection for all individuals over the time frame.

2.3. IMPLEMENTATION

We run the algorithm for 200,000 steps with four chains, with a 100,000 step burn-in and thinning of 1000, giving 200 posterior samples.

3. The model specification in *serojump* for the empirical data

3.1 DATA OVERVIEW

| **Variable** | **Values** |
| --- | --- |
| Number of individuals (i) | {1, 2, …, 256} |
| Length of study period (t) | {1, 2, …, 308} |
| Biomarkers measured (b) | Ancestral spike and NCP |
| Exposure types considered | Pre-infection ($e_{pre})$  Post-infection with pre-delta variant ($e_{pre-delta})$  Post-infection with delta variant ($e_{delta})$  Post-vaccination ($e_{vax}$) |
| Fitted exposure type | Post-infection with delta variant   - A) with no known infection - B) with Some known infections (from PCR) |

Table SM3. Overview of the serological data in the empirical dataset.

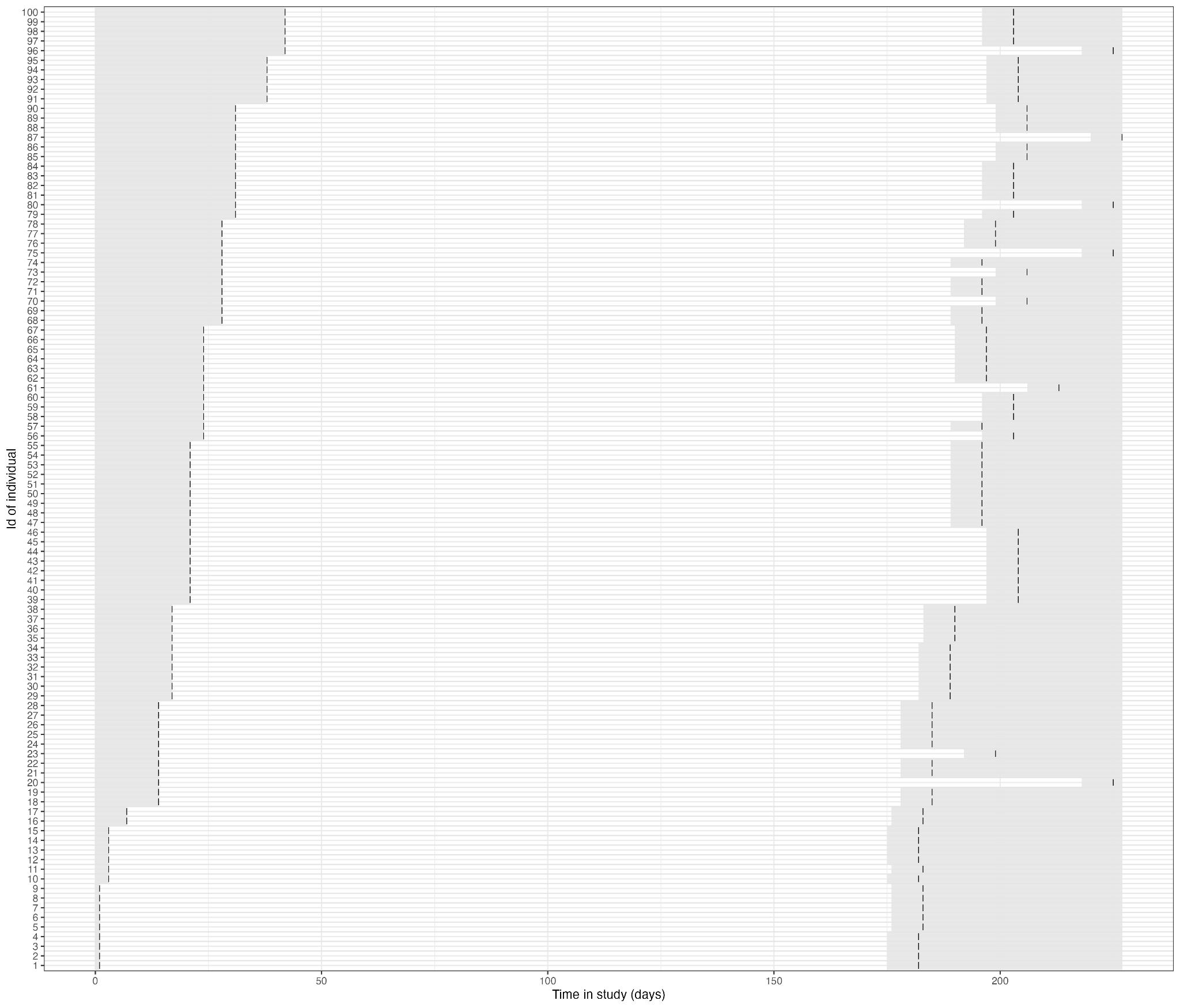

Figure SM4. **Longitudinal timeline of individual follow-up in the study.** Each row represents a unique individual (showing first 100), with the x-axis showing the time in the study (days). Gray bars indicate the period where an infection cannot be inferred, while black points mark dates that serological samples were taken.

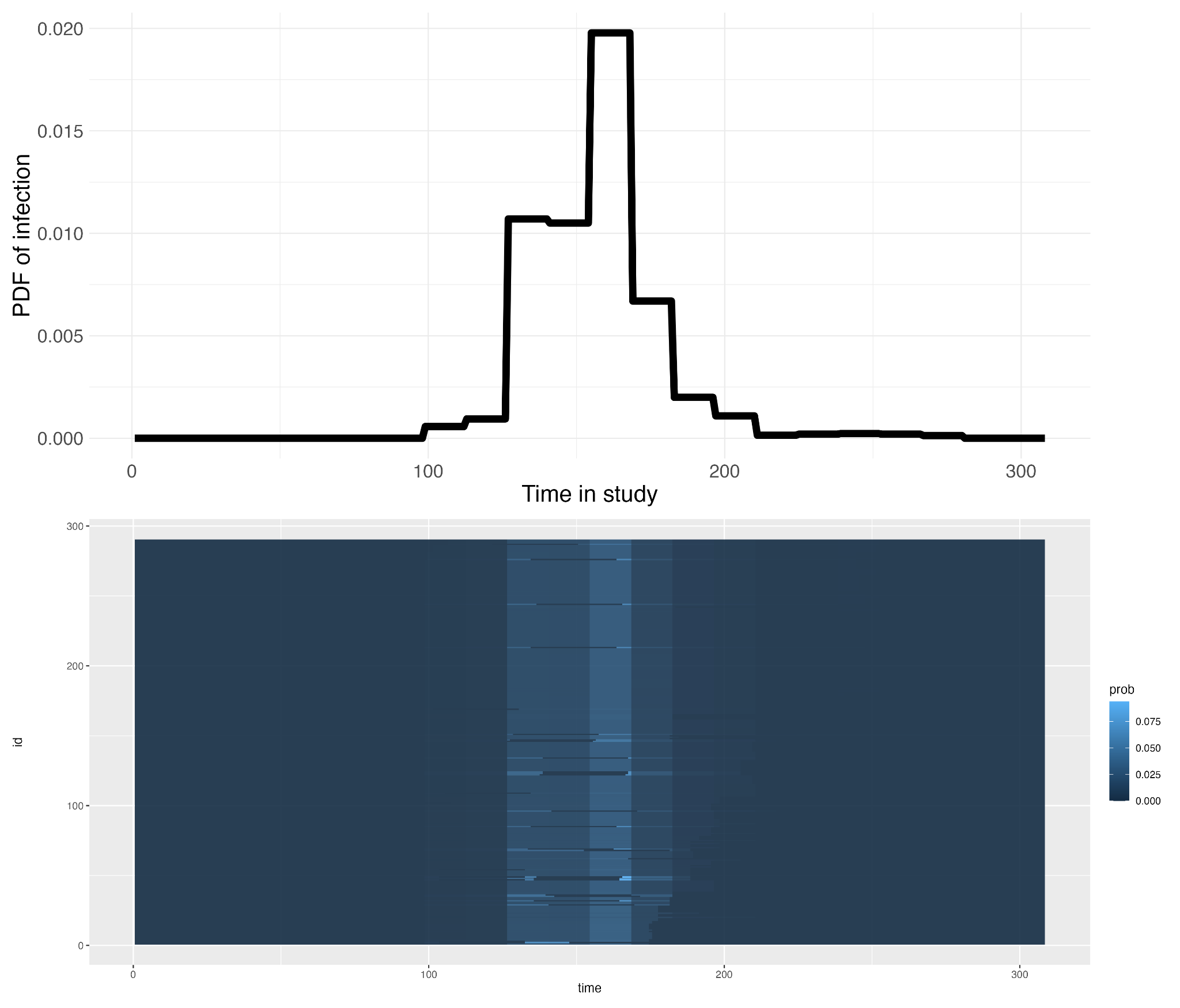

**Figure SM5. Probability density and individual-level posterior probabilities of infection over time.** The top panel shows the probability density function (PDF) of infection events across the study period (empirical probability density function). The x-axis represents time in the study (days), and the y-axis represents the PDF of infection. The bottom panel provides a heatmap of posterior probabilities of infection for each individual (y-axis) over time (x-axis). Lighter blue shades indicate higher probabilities of infection.

3.2. MODEL OVERVIEW (A + B)

| OBSERVATIONAL MODEL: $P_{obs}(Y_{i, t}^{b}\vert A_{i,t}^{b},\theta)$ | | |
| --- | --- | --- |
|  | Likelihood function: | P_obs_ = *f_N_*(A_i, t_^b^\| Y_i, t_^b^, 𝜎)  Priors: 𝜎 ~ Uniform(0, 4) |
| ANTIBODY KINETICS: $f_{e_{pre}}$,$f_{e_{pre-delta}}$,$f_{e_{delta}}$,$f_{e_{vax}}$ for Ancestral spike and NCP (see Figure SM6 for Prior Predictive distribution) | | |
|  | Pre-infection ($e_{pre}$) | **Functional** Form:$f_{e_{pre}}(t, Y_{i,0}) =Y_{i,0} - tw$  **Priors**: w ~ Uniform(0, 0.1) |
|  | Post-infection with pre-delta variant (  $e_{pre-delta})$  Post-infection with delta variant ($e_{delta})$  Post-vaccination ($e_{vax}$) | **Functional Form:**  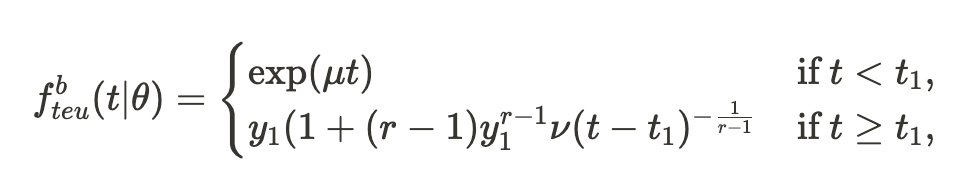  **Priors**: r ~ U(0, 1), y_1_ ~ U(0, 6), t_1_ ~ U(7, 21), v = 0.001  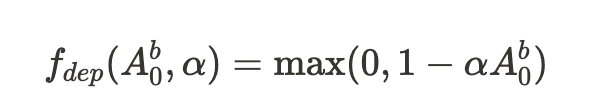  Priors: $\alpha$ ~U(0, 1) |
| PRIOR ON INFECTION TIME (*delta*), $P_{exp}(\tau_{i})$ | | |
|  | For each individual, *i* | See Figure SM5. We use the empirical distribution from the PCR cases. We also set a 0 probability of infection two weeks after a known pre-delta infection for an individual |
| PRIOR ON TOTAL NUMBER OF INFECTIONS, $P(Z)$ | | |
|  | For each individual , *i* | Z_i_ ~ Bernoulli(0.5) |

Table SM4. Overview of the model inputs fort *serojump* for the empirical dataset.

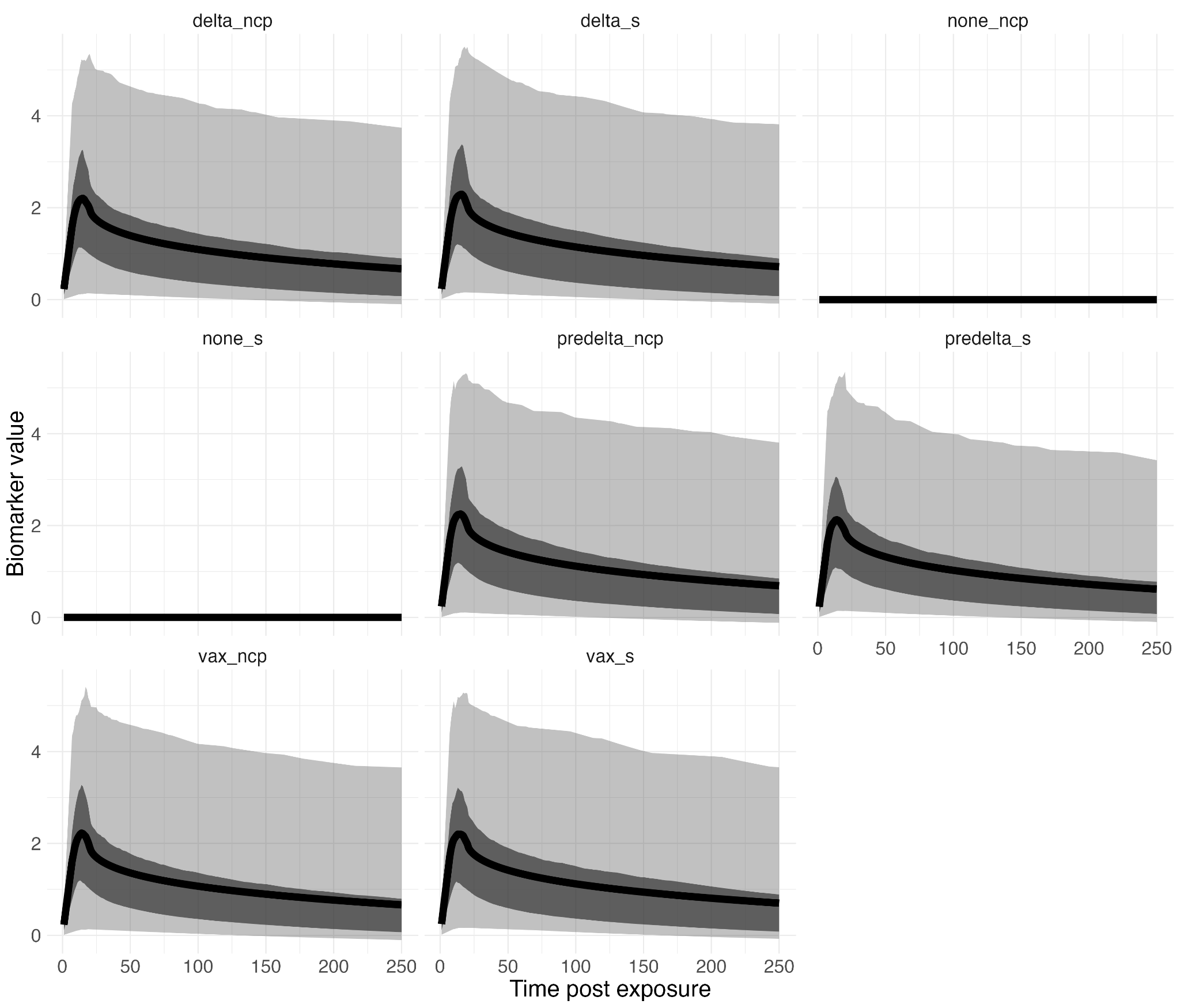

**Figure SM6. Prior predictive distributions for antibody kinetics.** The solid black line represents the median predicted antibody response over time, while the shaded area shows the uncertainty (e.g., 95% credible interval) in the prior predictions. The x-axis represents time (days), and the y-axis indicates the modeled antibody level. We show the distributions for infection with delta variant, pre-delta variant, pre-infection and vaccination for spike and NCP antigen.

3.3. IMPLEMENTATION

We run the algorithm for 200,000 steps with four chains, with a 100,000 step burn-in and thinning of 1000, giving 200 posterior samples.
