## Supplementary figures and images for "*serojump*: A Bayesian tool for inferring infection timing and antibody kinetics from longitudinal serological data"

### Figure S1

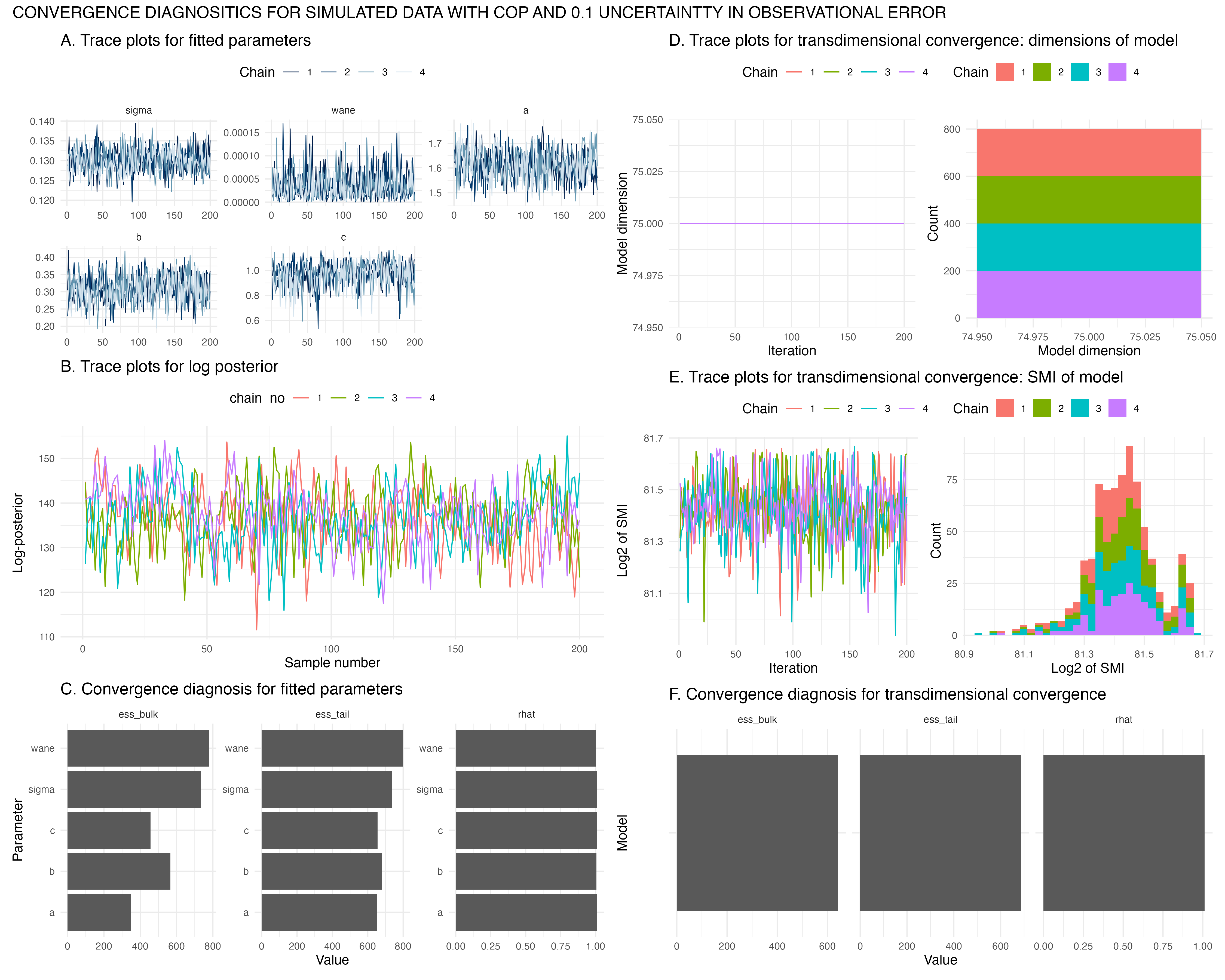

### Figure S2

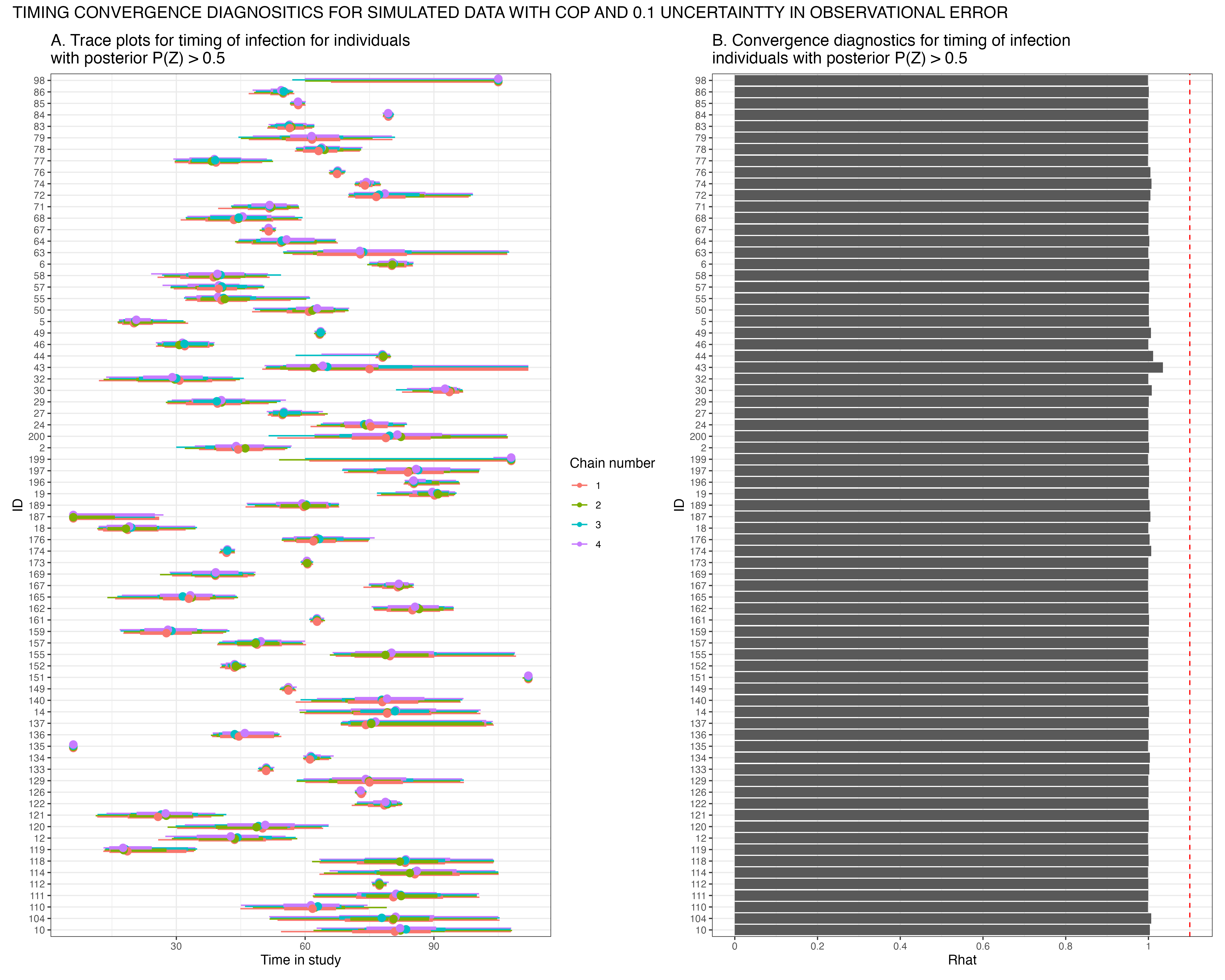

### Figure S3

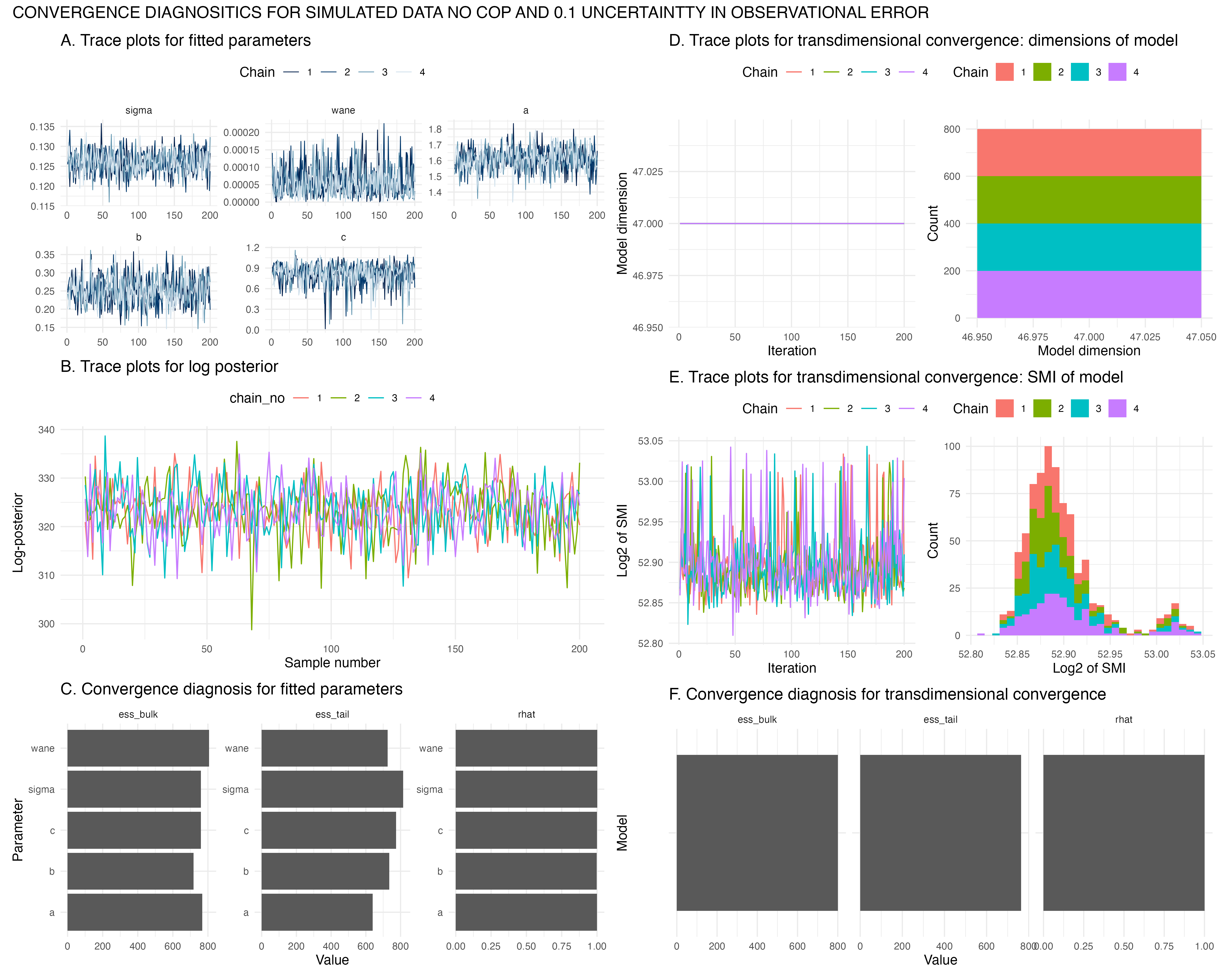

### Figure S4

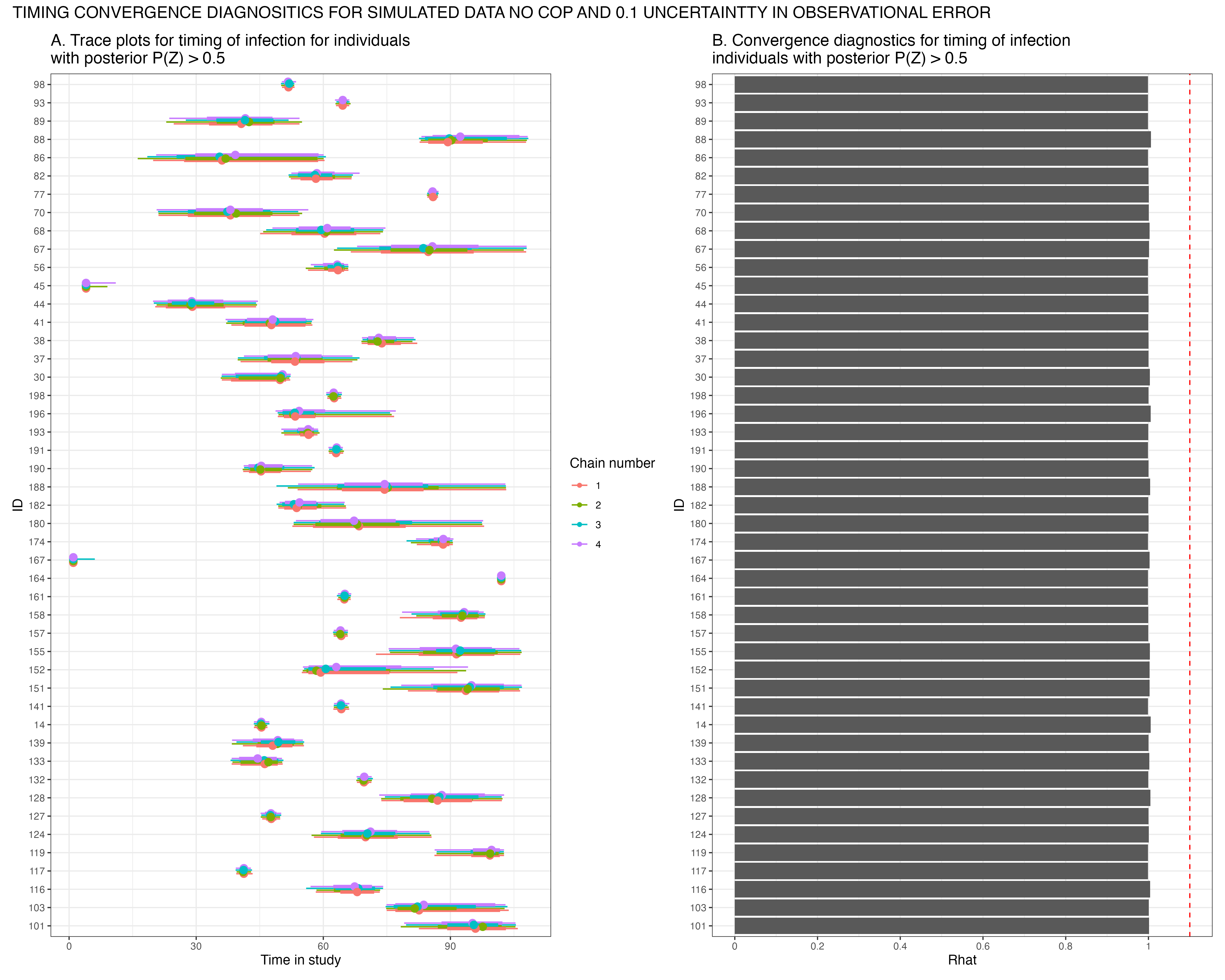

### Figure S5

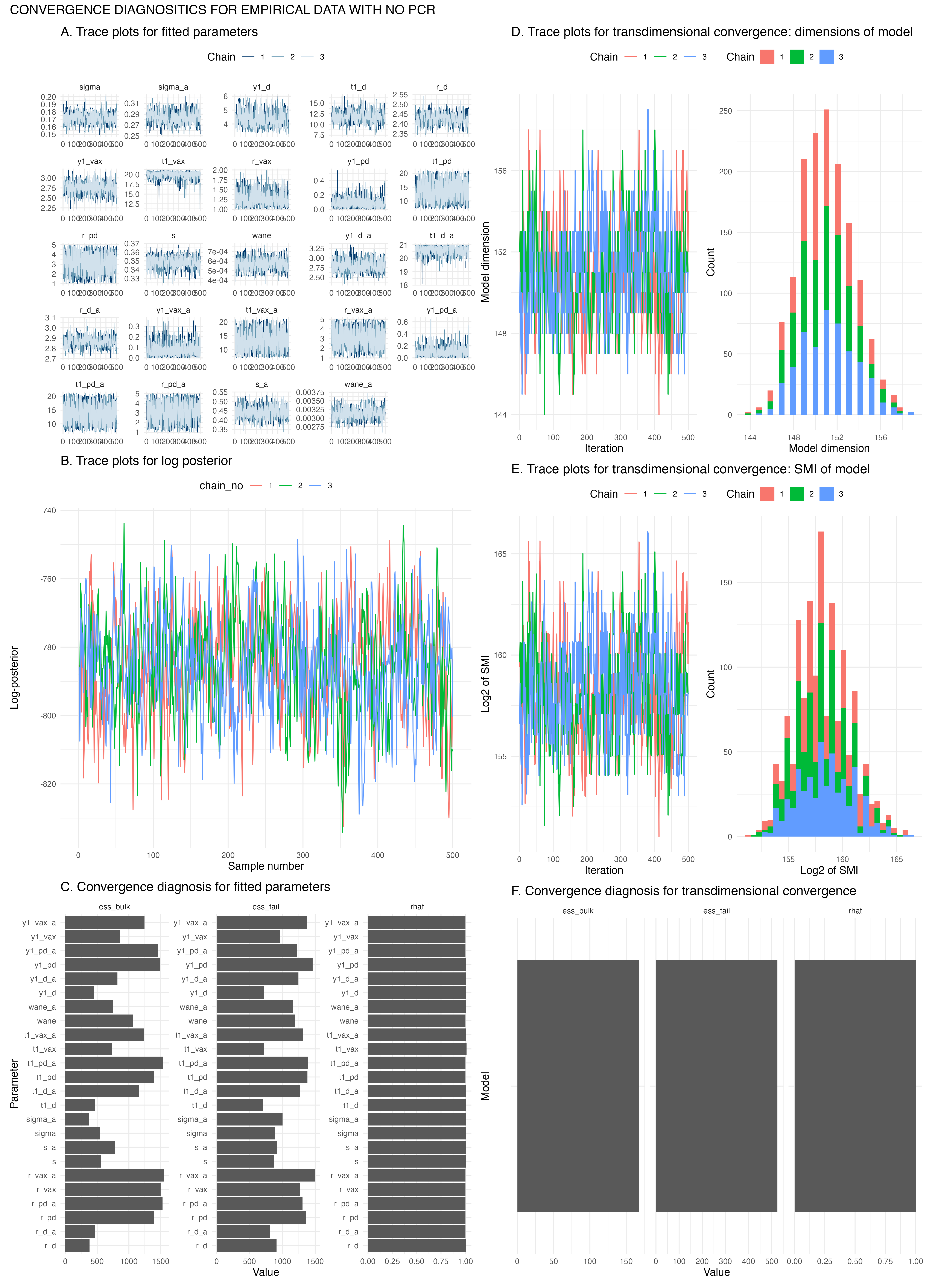

### Figure S6

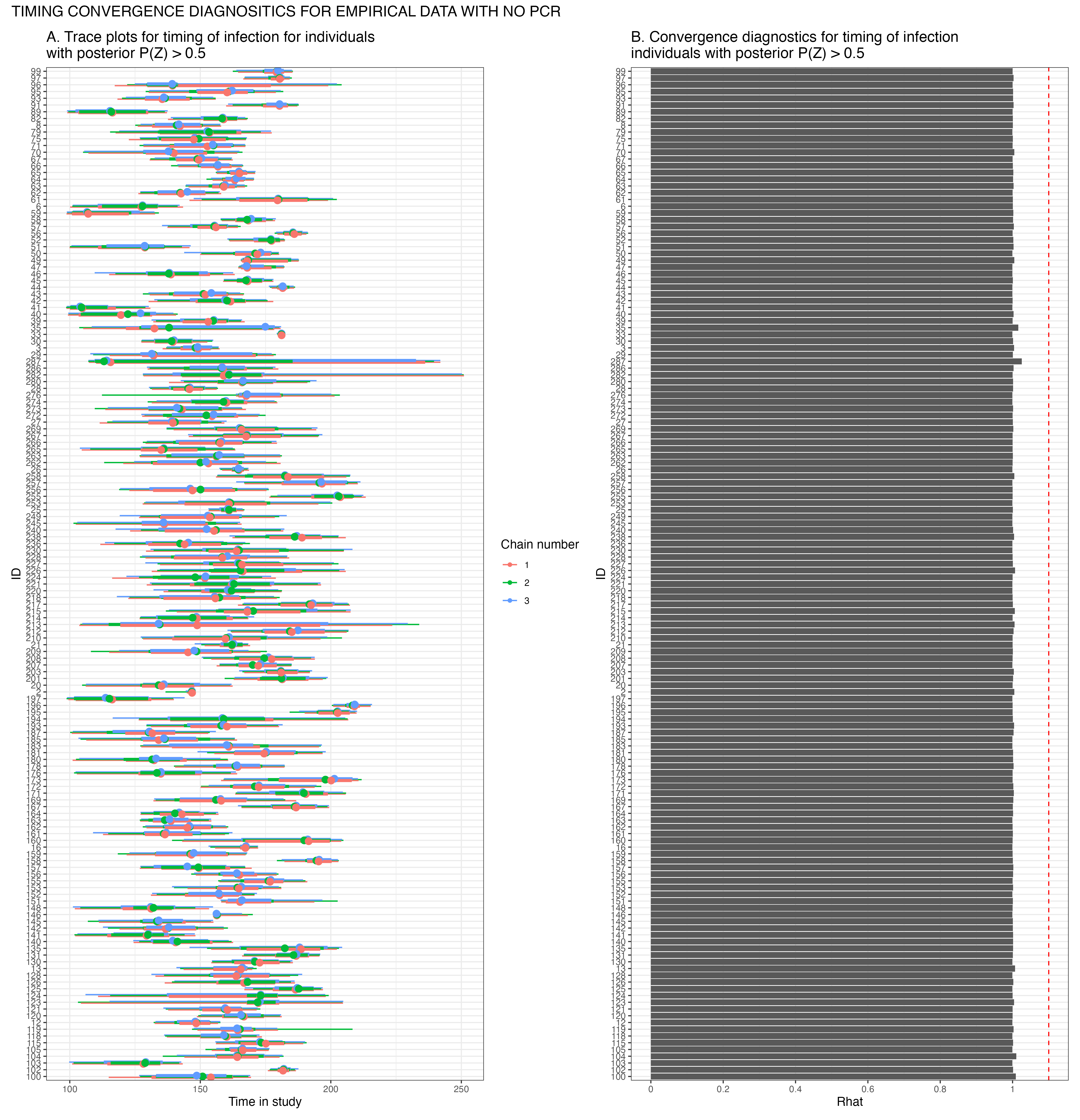

### Figure S7

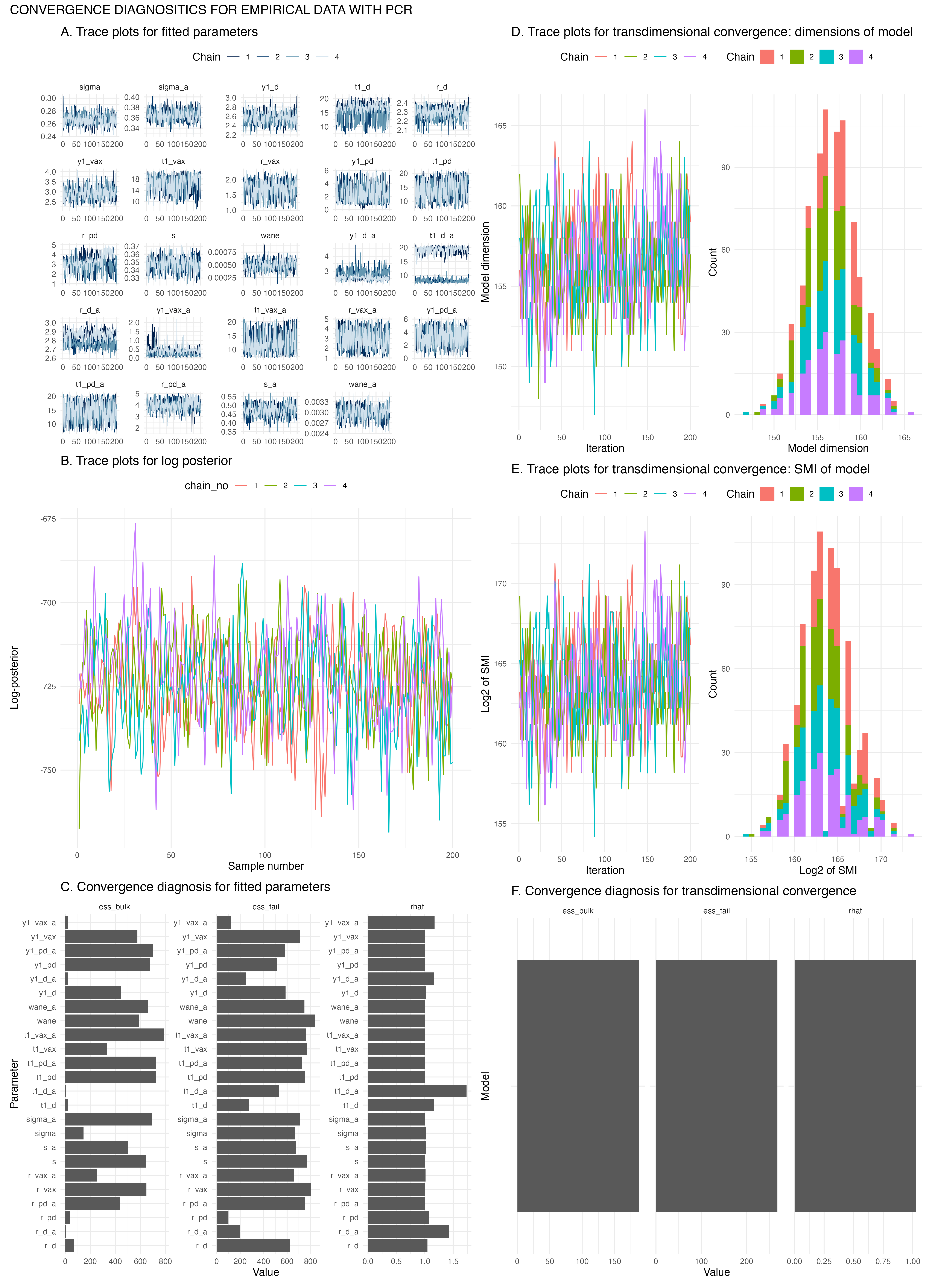

### Figure S8

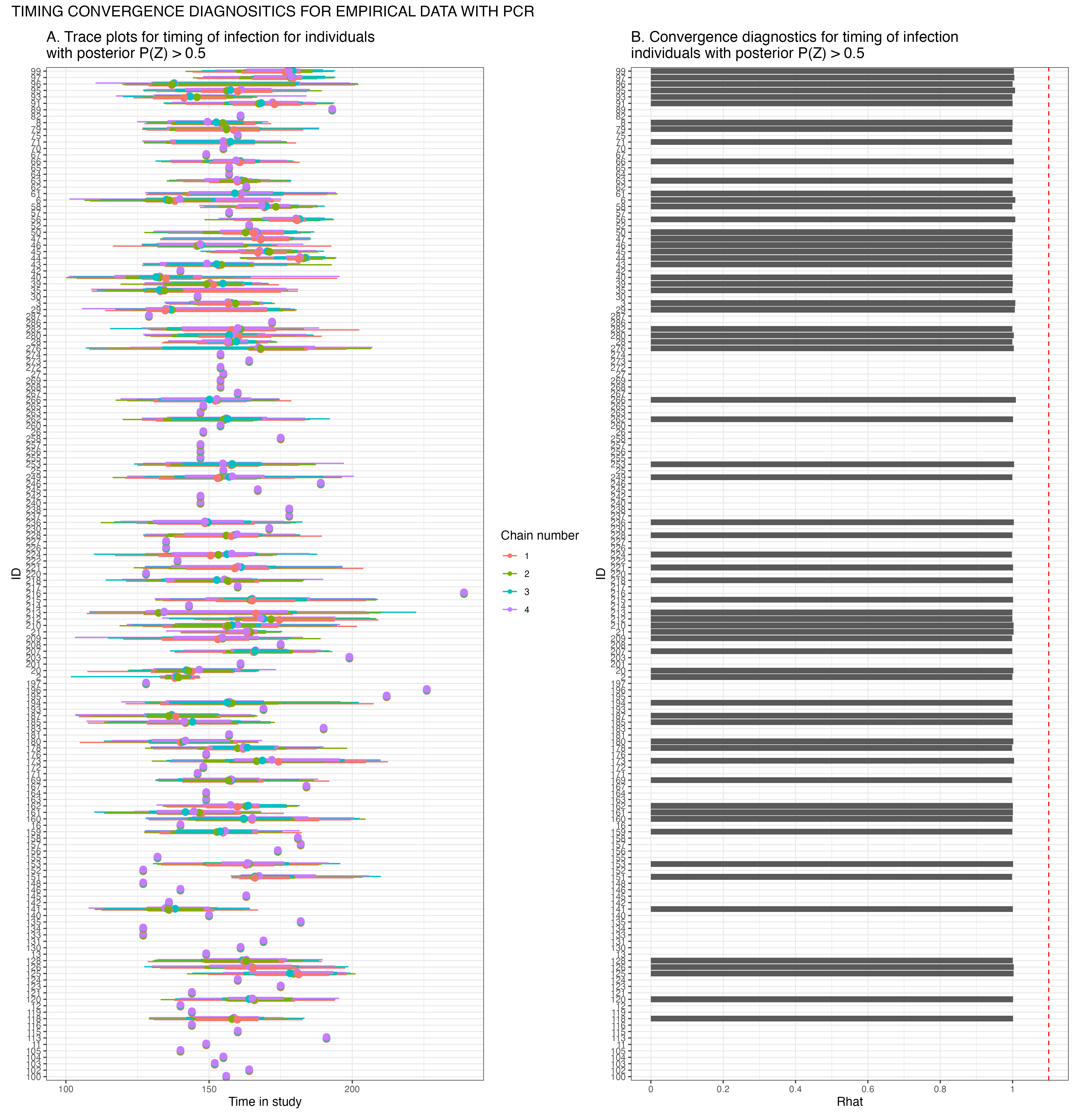

### Figure S9

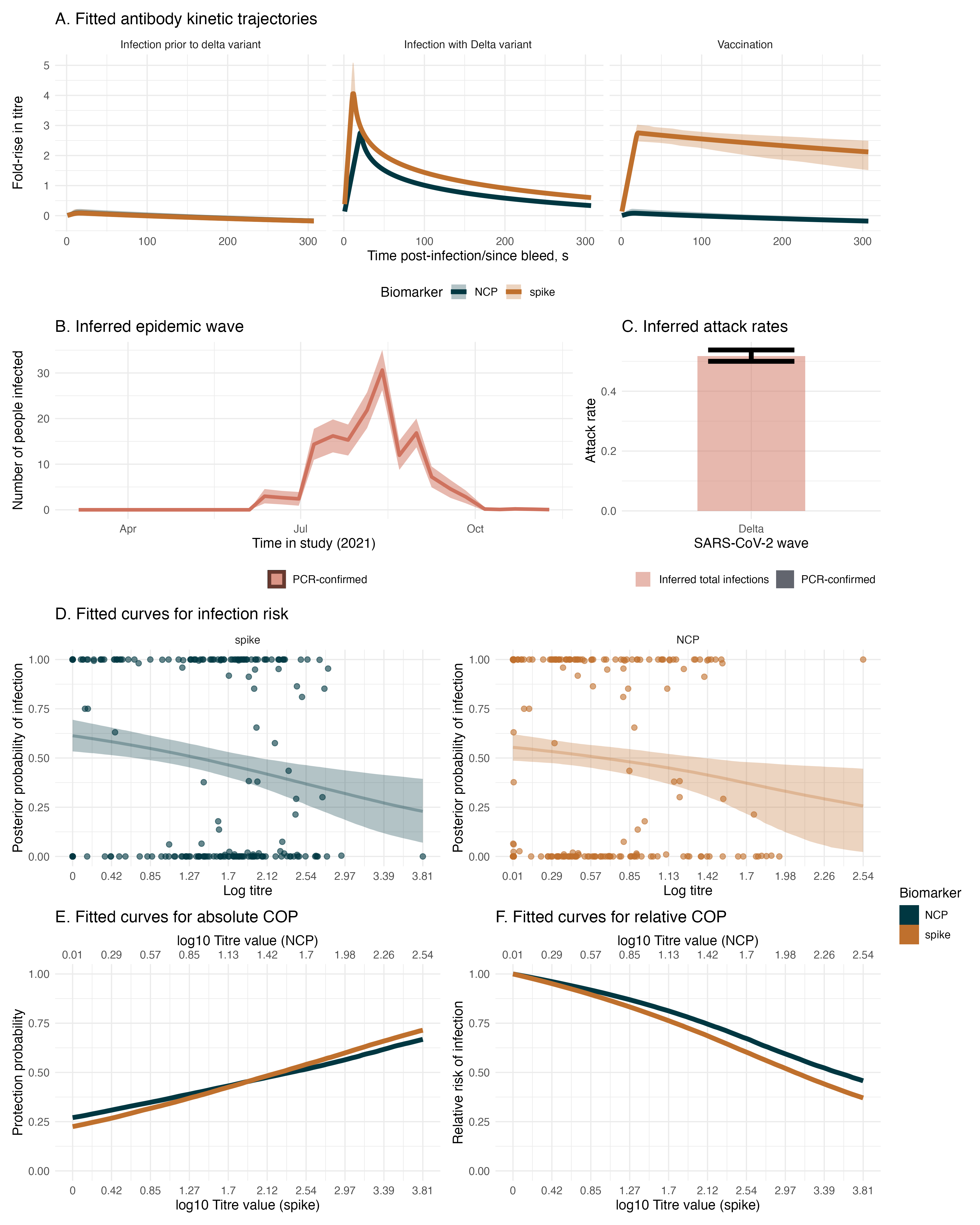
